# T cell-intrinsic lymphoproliferation as a key driver of intestinal autoimmunity in acquired generalized lipodystrophy

**DOI:** 10.1101/2025.02.19.25322160

**Authors:** Marilena Letizia, Toka Omar, Patrick Weidner, Manuel O. Jakob, Inka Freise, Susanne M. Krug, Britt-Sabina Löscher, Elisa Rosati, Benedikt Obermayer, Maria de los Reyes Gamez-Belmonte, Julia Hecker, Joern-Felix Ziegler, Benjamin Weixler, Patrick Asbach, Desiree Kunkel, Michael Stumvoll, Konstanze Miehle, Christoph Becker, Christoph S.N. Klose, Rainer Glauben, Dieter Beule, Anja Kühl, Andre Franke, Ashley Sanders, Britta Siegmund, Carl Weidinger

**Author notes:** These authors contributed equally to this work (Marilena Letizia, Toka Omar, Patrick Weidner). These authors jointly supervised this work and serve as corresponding authors (Britta Siegmund, Carl Weidinger).

## Abstract

Acquired generalized lipodystrophy (AGL) is a rare metabolic disorder frequently associated with autoimmunity. Its etiology is incompletely understood and the impact of adipose tissue loss on autoimmunity and intestinal inflammation in AGL remains unclear. Using mass cytometry and single-cell RNA sequencing, we observed an oligoclonal expansion of T cells in the periphery and inflamed intestine in a patient with AGL and Crohn’s disease (AGLCD). To explore if loss of adipose tissue triggers lymphoproliferation, we studied lipodystrophic mice as a model for AGL. Unexpectedly, lipodystrophic mice did not show T-cell expansion, were protected from colitis and displayed a defect in the development of pro-inflammatory T cells, which could be reversed by allogeneic fat transplantations, indicating that clonal T-cell expansion is not primarily caused by lipodystrophy. Instead, gene sequencing revealed a T cell-intrinsic de-novo *NRAS* mutation, pointing towards somatic mosaicism as a driver of clonal T-cell expansion and systemic autoimmunity in AGLCD.

## Introduction

Acquired generalized lipodystrophy (AGL) is a complex metabolic disorder of non-hereditary origin. It is characterized by the complete absence of adipose tissue, severe type 2 diabetes, and early onset of metabolic complications including the development of pronounced steatohepatitis^1, 2^. AGL is frequently associated with autoimmune comorbidities, including autoimmune hepatitis, arthritis, glomerulonephritis, and Crohn’s disease (CD). Additionally, AGL patients are at an elevated risk of developing rare forms of T-cell lymphoma^3^.

The etiology of AGL remains poorly understood. However, initial observations in melanoma patients suggest a T-cell-driven development of AGL since several cases of progressive loss of adipose tissue and AGL have been reported in cancer patients following immunomodulatory therapies with the checkpoint blockers nivolumab or pembrolizumab^4, 5, 6^. The high incidence of T-cell lymphoma occurring in AGL patients further supports the hypothesis that T cells may play a critical role in AGL pathogenesis^3^. Nevertheless, it remains unclear whether the increased incidence of autoimmune disorders in AGL patients directly results from the absence of fat-derived immunomodulatory signals or whether auto-inflammation is primarily caused by T cell-intrinsic disturbances of immune cell homeostasis. Additionally, the precise manner by which the absence of adipose tissue and missing fat-derived signals influences the onset of autoimmunity in AGL is not understood.

In addition to the clinical necessity of elucidating the pathophysiology of AGL, human lipodystrophies are of particular interest for metabolic research since they can serve as a powerful model system to study the interplay between adipose tissue and the immune system. To date, the role of adipose tissue in the pathophysiology of inflammatory bowel diseases (IBD) such as CD remains elusive. However, it is well established that patients with CD commonly exhibit specific alterations in the mesenteric fat, also referred to as “creeping fat” (CF). These changes include the development of hyperplastic adipocytes, increased local production of various adipokines including leptin and adiponectin, infiltration with lymphocytes and macrophages, as well as colonialization of CF with live bacteria^7, 8^. Furthermore, recent epidemiologic meta-analyses have indicated that there may be an association between obesity and an increased risk of developing CD^9^. There is also evidence that obesity is linked to a worse clinical outcome in CD patients, comprising an elevated risk of disease recurrence, therapeutic failure, and higher resection rates^10^. However, it remains challenging to decipher the causative contribution of individual adipose tissue-derived signals to the pathogenesis of IBD in humans.

By studying a patient with AGL and concomitant Crohn’s disease (AGLCD) we previously observed that treatment of the AGLCD patient with recombinant leptin altered immune cell function and metabolism of macrophages as well as T cells. Thus, leptin substitution induced TNFα in T cells and monocytes *in vivo* and worsened intestinal autoimmunity in the AGLCD patient, suggesting that leptin acts as a proinflammatory adipokine in human intestinal inflammation^11^. Similarly, previous data indicate that leptin receptor (LEPR)-deficient CD4^+^ T cells of *Lepr^fl/fl^-Cd4-Cre* mice fail to induce intestinal and neuronal inflammation in T cell transfer models^12, 13^ as LEPR-deficient mice show impaired Th17 cell differentiation *in vitro* and *in vivo*^13^, underscoring leptin’s proinflammatory role in IBD pathogenesis.

In this study, we aimed to elucidate the molecular mechanisms underlying autoimmunity in AGL. We characterized a rare patient with AGLCD and used lipodystrophic *Pparg^fl/fl^-Adipoq-Cre* mice^14^ to explore the role of adipose tissue in intestinal autoimmunity under steady-state conditions, following DSS-induced colitis, and through allogeneic fat transplantation models. Mass cytometry and scRNA sequencing revealed an AGLCD-specific oligoclonal expansion of T cells associated with a somatic *NRASG13D* mutation, previously linked to autoimmune lymphoproliferative syndrome^15^, suggesting that somatic mosaicism might contribute to clonal T cell expansion in AGL. By using mouse models of lipodystrophy, we could observe that adipose tissue loss does not primarily cause T cell lymphoproliferation; instead, lipodystrophic mice were protected from DSS-induced colitis due to impaired Th1 and Th17 cell function, which could be partially reversed by allogenic transplantation of wild-type (WT) fat but not of leptin-deficient adipose tissue. Accordingly, treatment of the AGLCD patient with recombinant leptin increased the abundance of Th17 cells *in vivo*, underscoring leptin’s role as a key regulator of pro-inflammatory T cells. Overall, we present evidence that AGLCD resembles a clonal autoimmune lymphoproliferative disease characterized by an oligoclonal expansion and a T cell-intrinsic *de novo NRASG13D* mutation. Furthermore, our findings indicate that reducing adipose tissue mass may be beneficial to modulate intestinal inflammation in IBD by suppressing adipokine-induced pro-inflammatory T cells.

## Results

### Detection of high abundances of effector memory CD4^+^ and CD8^+^ T cells in the periphery and inflamed intestine of an AGLCD patient

The pathogenesis of AGL remains poorly understood, and the drivers of autoimmunity in AGL have yet to be elucidated. To gain insights into the mechanisms of autoimmunity in AGL, we conducted an in-depth analysis of the intestinal immune cell composition of a rare patient with AGLCD^11^, who suffered from a complete absence of adipose tissue, concomitant metabolic dysfunction-associated fatty liver disease and severe ileal and colonic inflammation (**Figure 1A**). Thus, we compared the AGLCD patient to fat-proficient non-inflamed controls or patients with CD by mass cytometry (**Figure 1B**, **Suppl. Figure 1**) resolving a total of 17 intestinal immune cell clusters (**Figure 1C-D**). Among these, CD4^+^ IL-7R^-^ CD25^+^ regulatory (cluster 8) as well as CD4^+^ CD45RO^+^ effector (cluster 9) T cells, which have been identified as a cell signature indicative of inflammatory bowel disease (IBD)^16^, were enriched in both the AGLCD and CD patients when compared to non-inflamed controls (**Figure 1D**). Interestingly, only the AGLCD patient showed an expansion of CD8^+^ CD45RO^+^ effector T cells (cluster 13), which was not detected in CD patients or non-inflamed controls (**Figure 1E-F**). **Suppl. Figure 2** summarizes the differences in expression levels of activation markers, cytokines, and pore-forming cytolytic proteins between cell clusters of the AGLCD patient, CD patients, and non-inflamed controls.

**Figure 1.**
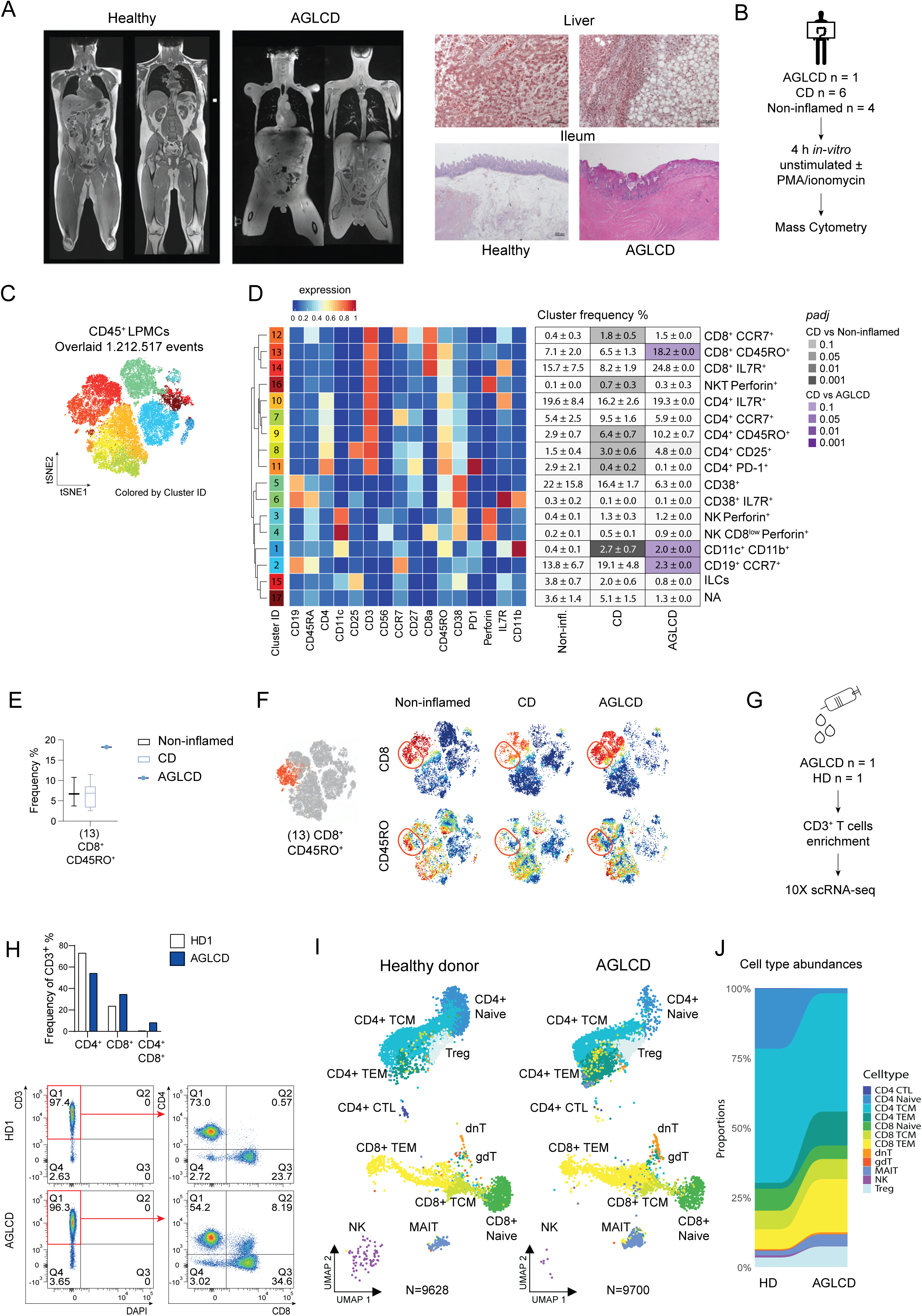
Enrichment of CD8^+^ T cells in the periphery and small intestinal lamina propria in a patient with acquired generalized lipodystrophy and combined Crohn’s disease (AGLCD). **(A)** T1-weighted whole-body MRI scan (left) and histologic liver (upper right) and gut sections (lower right) of a rare patient with acquired generalized lipodystrophy combined with Crohn’s disease (AGLCD) compared to a male healthy individual showing the complete absence of fat tissue in the AGLCD patient, as well as steatohepatitis and severe ileal inflammation. **(B)** Experimental setup for mass cytometry. Lamina propria mononuclear cells (LPMCs) were isolated from terminal ileum samples obtained from 6 Crohn’s disease (CD) patients, one AGLCD patient, and 4 non-inflamed controls. Cells were activated *in vitro* for 4 h ± PMA/ionomycin and stored at −80°C until acquisition. **(C)** FlowSOM plot of merged FCS files. Colors indicate 17 clusters among CD45^+^ LPMCs. **(D)** Heatmap clusters show the expression levels of the 16 markers used for cluster analysis. On the right, the table shows the mean values (± SEM) of quantified frequencies (%) in unstimulated non-inflamed, CD, and AGLCD samples. Statistical differences were calculated using the edgeR statistical framework with negative binomial GLM and a false discovery rate adjusted to 10% using the Benjamini-Hochberg procedure. **(E)** Boxes show the frequency of CD8^+^ CD45RO^+^ effector T cells (cluster 13) in non-inflamed controls, CD patients, and the AGLCD patient. Boxes range from the 25th to the 75th percentiles. Whisker plots show the min (smallest) and max (largest) values. The line in the box indicates the median. **(F)** viSNE plots of an exemplary non-inflamed, a CD, and the AGLCD patient colored by marker expression levels (blue: low, red: high). Data in C-F are a summary of one mass cytometry experiment. **(G)** Experimental setup for scRNA-seq performed on CD3^+^ T cells isolated from the AGLCD patient and a healthy donor (HD). **(H)** The bar graphs quantify CD4^+^ and CD8^+^ T cell frequencies. FACS plots show the purity of CD3^+^ T cell enrichment (AGLCD n=1; HD n=1). **(I)** Umaps show the different cell clusters identified by scRNA-seq analysis in the AGLCD patient or a healthy individual (J) Bars show the frequency of cells per cluster. Data in G-J were obtained from one scRNA-seq experiment.

As observed in the intestinal lamina propria (LP), we had previously identified a systemic expansion of CD8^+^ T cells in the peripheral blood of the AGLCD patient^11^, which indicates that the AGLCD patient exhibits a persisting systemic occurrence of expanded CD8^+^ T cells. Therefore, to further investigate T cell functions in the AGLCD patient, we performed single-cell RNA sequencing (scRNA-seq) on 10,000 peripheral blood sort-purified CD3^+^ cells isolated from the AGLCD patient and a healthy donor (HD) (**Figure 1G-H**). scRNA-seq data were deconvoluted into 14 immune cell subsets (**Figure 1I**), revealing a high abundance of peripheral CD4^+^ and CD8^+^ effector memory T cell subsets, MAIT cells, and a reduced frequency of CD3^+^ NK cells in the AGLCD patient (**Figure 1J**). These findings confirm our previous observations of a defect in NK cell homeostasis in the AGLCD patient and the persisting expansion of CD8^+^ T cells over time^11^.

Given that adipose tissue plays a critical role in regulating the immune system in the context of IBD as well as rheumatoid arthritis^7, 17^ and given that AGL patients frequently experience autoimmune disorders such as arthritis, glomerulonephritis, and autoimmune hepatitis^2^, we sought to determine whether the observed accumulation of CD8^+^ T cells was a consequence of the absence of adipose tissue. Accordingly, we examined the impact of adipose tissue loss on immune cell composition, focusing on T cell differentiation, function, and expansion in lipodystrophic *Pparg^fl/fl^-Adipoq-Cre* mice.

### Lipodystrophic mice show alterations in splenic NK and intestinal T cell composition and have increased intestinal antimicrobial peptide expression

As previously published by Wang *et al.*,^14^ lipodystrophic *Pparg^fl/fl^-Adipoq-Cre* (Lipo) mice exhibited a complete absence of visceral fat (**Figure 2A**), consecutive absence of serum levels of adipokines including leptin (**Figure 2B**) and developed non-alcoholic steatohepatitis analogous to our AGLCD patient (**Figure 2C, Suppl. Figure 3A**). However, no accumulation of lipids was observed in other tissues, including the pancreas, spleen, kidneys, or cardiac muscle (**Suppl. Figure 3A**). Additionally, splenomegaly with an enlarged red bulb and larger-sized glomeruli in the kidneys were also detected, as previously described^18^ (**Suppl. Figure 3A**). Although it has been reported that knockout models of fat-derived signals, such as leptin (*ob/ob*) or leptin receptor-deficient (*db/db*) mice exhibit disturbances in immune cell homeostasis compared to WT controls^19^, scarce knowledge exists regarding the impact of lipodystrophy on immune cell homeostasis, and more specifically on T cell homeostasis. To gain insight into the impact of adipose tissue loss on immune cell composition in lipodystrophic mice at steady state, we conducted a comprehensive immune cell characterization of the spleen and intestinal tissue using flow cytometry (gating strategies are summarized in **Suppl. Figure 3B-C**). Specifically, following the isolation, splenocytes and lamina propria mononuclear cells (LPMCs) isolated from the ileum and colon were stained with antibody cocktails targeting CD4 and CD8 for the identification of T cells, CD19 for B cells, and Gr-1 for myeloid subsets. NK1.1, CD49b, CD49a, T-bet, Eomes, CD11b, and CD27 were used to characterize NK subsets according to the 4-stage model of NK cell maturation CD11b^low^ CD27^low^ → CD11b^low^ CD27^high^ → CD11b^high^ CD27^high^ → CD11b^high^ CD27^low 20^. Moreover, to identify discrepancies in cytokine production, cells were stimulated *in vitro* with PMA/ionomycin for 4 h and then stained intracellularly for TNFα, IFNγ, and IL-17A. Notably, lipodystrophic mice showed a diminished frequency of splenic NK1.1^+^ NK/ILC1 cells and an increased abundance of Gr1^+^ myeloid subsets in comparison to WT littermate controls (**Figure 2D-E**). Further analysis of the CD49b^+^ NK cell population revealed a higher frequency of less mature CD11b^high^ CD27^high^ NK cells and a reduced percentage of the highly cytolytic CD11b^high^ CD27^low^ NK subset in lipodystrophic mice compared to WT mice (**Figure 2F-G**). In addition, although no histological changes were detected in small and large intestinal tissue of lipodystrophic and WT animals (**Figure 2H**), flow cytometric analysis revealed a significant decrease in IL-17-producing CD4^+^ T cells in the small intestine (**Figure 2I-J**), as well as a significant reduction of CD8^+^ T and CD4^+^ regulatory T cells (Tregs) in the colon (**Figure 2K-L**) in fat-deficient lipodystrophic mice compared to WT controls.

**Figure 2.**
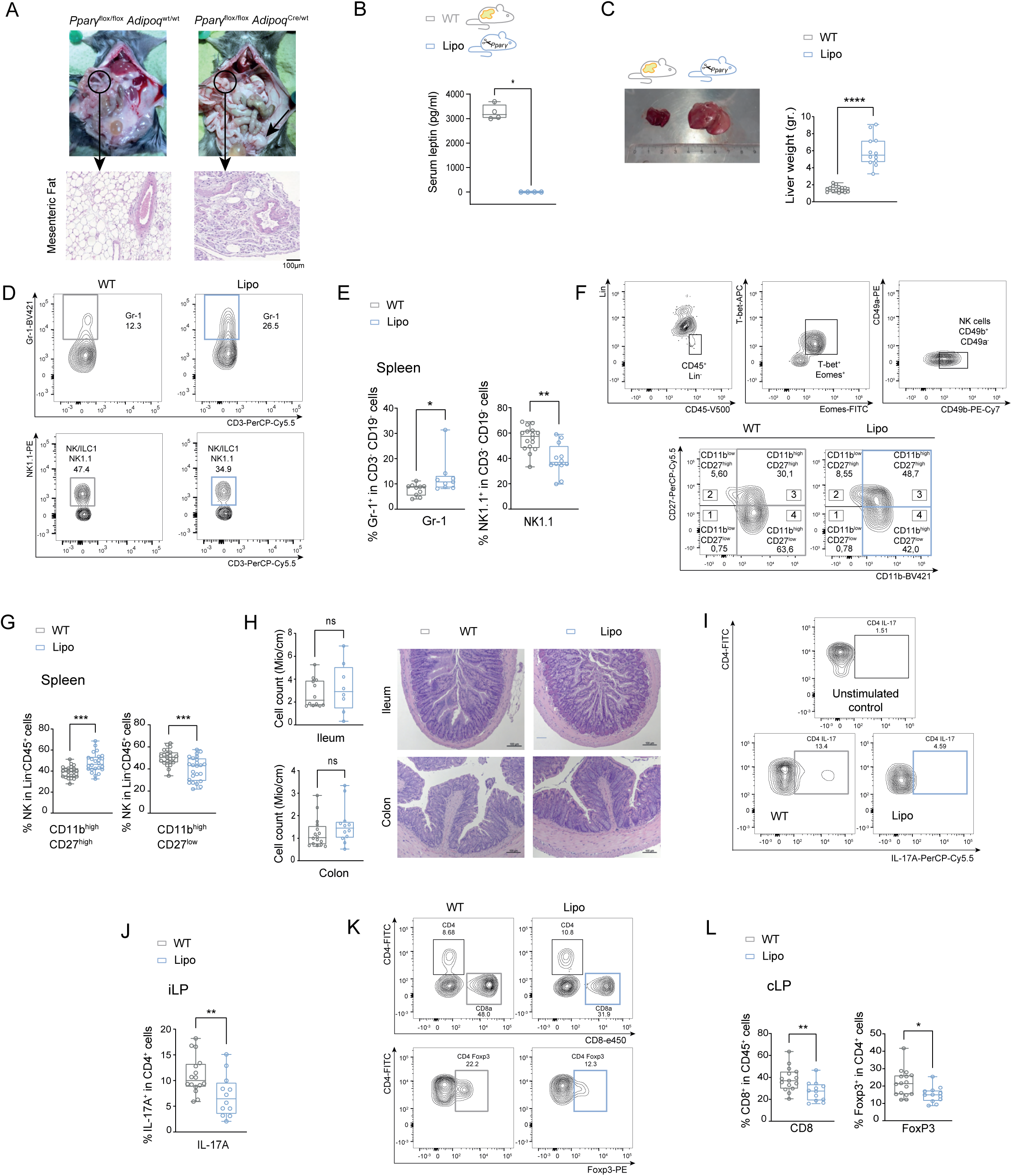
Lipodystrophic *Pparg^fl/fl^-Adipoq-Cre* mice mimic the hepatic phenotype of the AGLCD patient and show defects in the development of NK- and T cells under steady-state conditions. (**A**) Representative images of *Pparg^fl/fl^-Adipoq^Cre^* (Lipo) and wild-type (WT) mice showing the complete absence of fat tissue. Bottom, H&E staining of mesenteric adipose tissue (arrows indicate mesenteric and gonadal fat tissue). (**B**) Leptin serum levels (WT n=4; Lipo n=4). (**C**) Liver size of WT (n=16) or Lipo (n=12) mice from two independent experiments with exemplary pictures. (**D-G**) Flow cytometric assessment of splenic NK cells in Lipo mice (n=9-12) compared to WT littermates (n=10-16). Pooled data from 3 independent experiments. (**H**) Representative H&E staining and immune cells count in millions (Mio) per cm in the terminal ileum or colon as assessed by histology (WT: n=16; Lipo: n=12). (**I-J**) Gating strategy and frequency of CD4^+^ IL-17^+^ isolated from ileum lamina propria (iLP); (WT: n=16; Lipo: n=12). (**K-L**) Frequency of CD8^+^ and CD4^+^ Foxp3^+^ cells isolated from the colonic lamina propria (cLP); (WT: n=15-16; Lipo: n=12). Data was pooled from two independent experiments. All boxes range from the 25th to the 75th percentile. Whisker plots show the minimum (smallest) and maximum (largest) values. The line in the box indicates the median. Statistical differences in panel **B** were calculated using the Mann-Whitney U test, and in panels **C, E, G, H, J** and **L** using the unpaired t-test with Welch’s correction. \*\*\*\**P* < 0.0001, \*\*\**P* < 0.001, \*\**P* < 0.01, \**P* < 0.05.

These data collectively indicate that adipose tissue and its secreted factors play a pivotal role in maintaining immune cell homeostasis, including the differentiation of NK cells and intestinal T cells. Additionally, our findings show that the absence of adipose tissue and missing fat-derived signals in AGL are not responsible for the expansion of peripheral and intestinal CD4^+^ and CD8^+^ T cells observed in the AGLCD patient.

To gain further insight into the impact of adipose tissue absence on mucosal homeostasis, we conducted RNA bulk sequencing analyses of colonic tissue from 6 lipodystrophic and 6 control mice (**Figure 3A**). We detected 73 down-regulated and 83 up-regulated genes in the colonic tissue of lipodystrophic mice compared to WT littermates. Consistent with the complete absence of adipose tissue, the most significantly down-regulated genes in lipodystrophic mice were those transcribing for adipose-derived factors including adipsin (*Cfd*), resistin (*Retn*), leptin (*Lep*), and adiponectin (*Adipoq*) (**Figure 3B-C**). In addition, colonic tissue from lipodystrophic mice showed significantly higher mRNA transcripts of antimicrobial peptides (AMPs), including *Reg3b*, *Reg3g*, and *Chitin1*, compared to WT controls (**Figure 3D**). Additional pathway analyses revealed that lipodystrophic mice exhibited a differential regulation of genes primarily involved in adipocyte differentiation, regulation of thermogenesis, and pathways controlling lipid droplet formation (**Figure 3E-F**). Notably, we did not detect major alterations in intestinal epithelial cell differentiation between lipodystrophic and WT mice by RNA sequencing or qPCR (**Figure 3G**). Similarly, immunohistochemical staining of colonic tissue revealed comparable levels of chromogranin A and MUC2 expression, indicating that lipodystrophy does not affect the homeostasis of intestinal neuroendocrine and goblet cells (**Figure 3H**).

**Figure 3.**
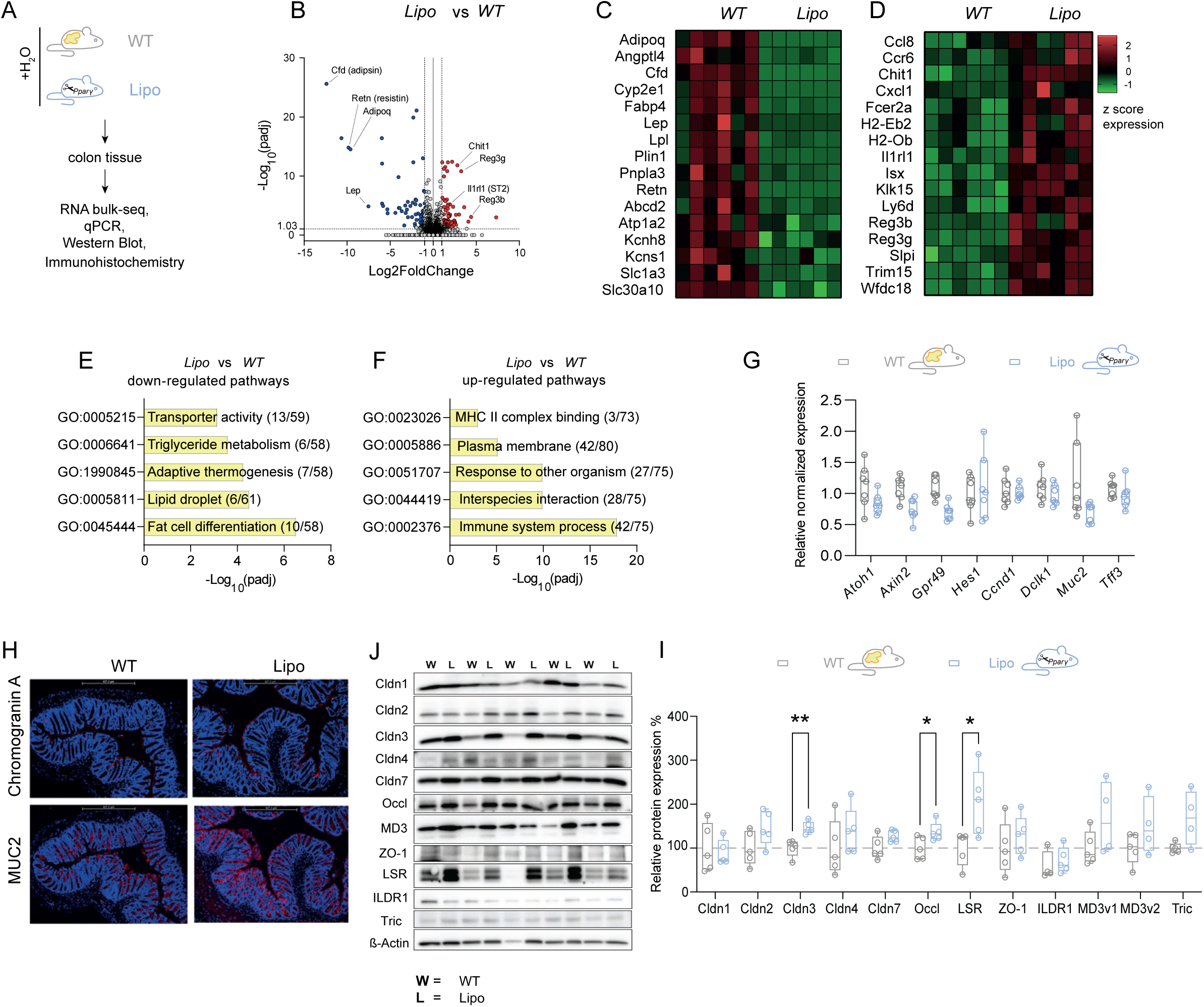
**Lipodystrophic mice display normal mucus production and harbor an increased expression of epithelial tight junction proteins**. (**A**) Experimental setup of RNA bulk sequencing experiments of colon tissue obtained from either wild-type (WT) or lipodystrophic (Lipo) mice. (**B**) Volcano plot showing differently regulated genes between Lipo and WT mice (n=6 per group). (**C-D**) Heatmaps showing differently regulated genes related to adipose tissue secreted factors and antimicrobial factors between Lipo mice and WT littermates (n=6 per group). (**E-F**) Bar graphs displaying pathway analyses of significantly regulated genes in lipodystrophic mice. (**G**) Box-and-whisker plots showing qPCR expression of epithelial differentiation markers in colonic tissue obtained from 6 WT or 6 Lipo mice. (**H**) Representative immunohistochemistry staining of colon tissue obtained from WT controls or Lipo mice showing epithelial MUC2 and chromogranin A expression. (**I-J**) Western blot analyses of colonic tight junction proteins in WT (n=5) and Lipo mice (n=5). Statistical differences in panels **G** and **J** were calculated using the Mann-Whitney U test. Boxes range from the 25th to the 75th percentile. Whisker plots show the minimum (smallest) and maximum (largest) values. The line in the box indicates the median. (SEM). \**P* < 0.05, \*\**P* < 0.01.

Several publications have proposed that excessive production of adipose-derived factors, such as leptin, could impair epithelial barrier function in obese patients by affecting the expression of tight junction proteins, including claudins, in intestinal epithelial cells^21, 22, 23^, which are crucial for epithelial barrier function and repair^24, 25^. We therefore proceeded to examine the impact of fat loss on tight junction (TJ) protein expression levels in intestinal epithelium cells by western blot analyses. Specifically, we analyzed members of the claudin (Cldn) family, which are expressed in the colon and mainly determine the paracellular barrier properties for ions and small solutes^26^. We furthermore studied TJ-associated MARVEL proteins (occludin, tricellulin, marvelD3), of which occludin and tricellulin are pivotal for paracellular macromolecule permeability, as well as the intestinal angulins angulin1/LSR and angulin2/ILDR1 which are controlling the correct localization of tricellulin and the regulation of the permeability of the tricellular TJ^23, 27^. As an important TJ-scaffolding protein, we additionally analyzed the expression of zonula occludens-1^26, 28^. As shown in **Figure 3J-I**, a significantly elevated expression of claudin 3 (Cldn3), occludin (Occl), and lipolysis-stimulated lipoprotein receptor (LSR) was observed in colonic tissue of lipodystrophic mice when compared to WT controls. This suggests that lipodystrophic mice may exhibit an augmented epithelial resistance to the translocation of luminal pathogens due to their enhanced expression of TJ proteins and antimicrobial peptides.

### Lipodystrophic mice are protected from chronic DSS-induced colitis

To rule out that the T lymphoproliferation observed in the AGLCD only arises during chronic intestinal inflammation in AGL, we next challenged lipodystrophic or WT littermates with three cycles of 1.5% DSS, which induced weight loss in both WT and lipodystrophic mice (**Figure 4A**). However, only fat-proficient WT mice developed severe chronic colitis with higher scores in stool consistency (**Suppl. Figure 4A**). In contrast, lipodystrophic animals were characterized by significantly reduced intestinal inflammation as assessed by histological analyses (**Figure 4B**).

**Figure 4.**
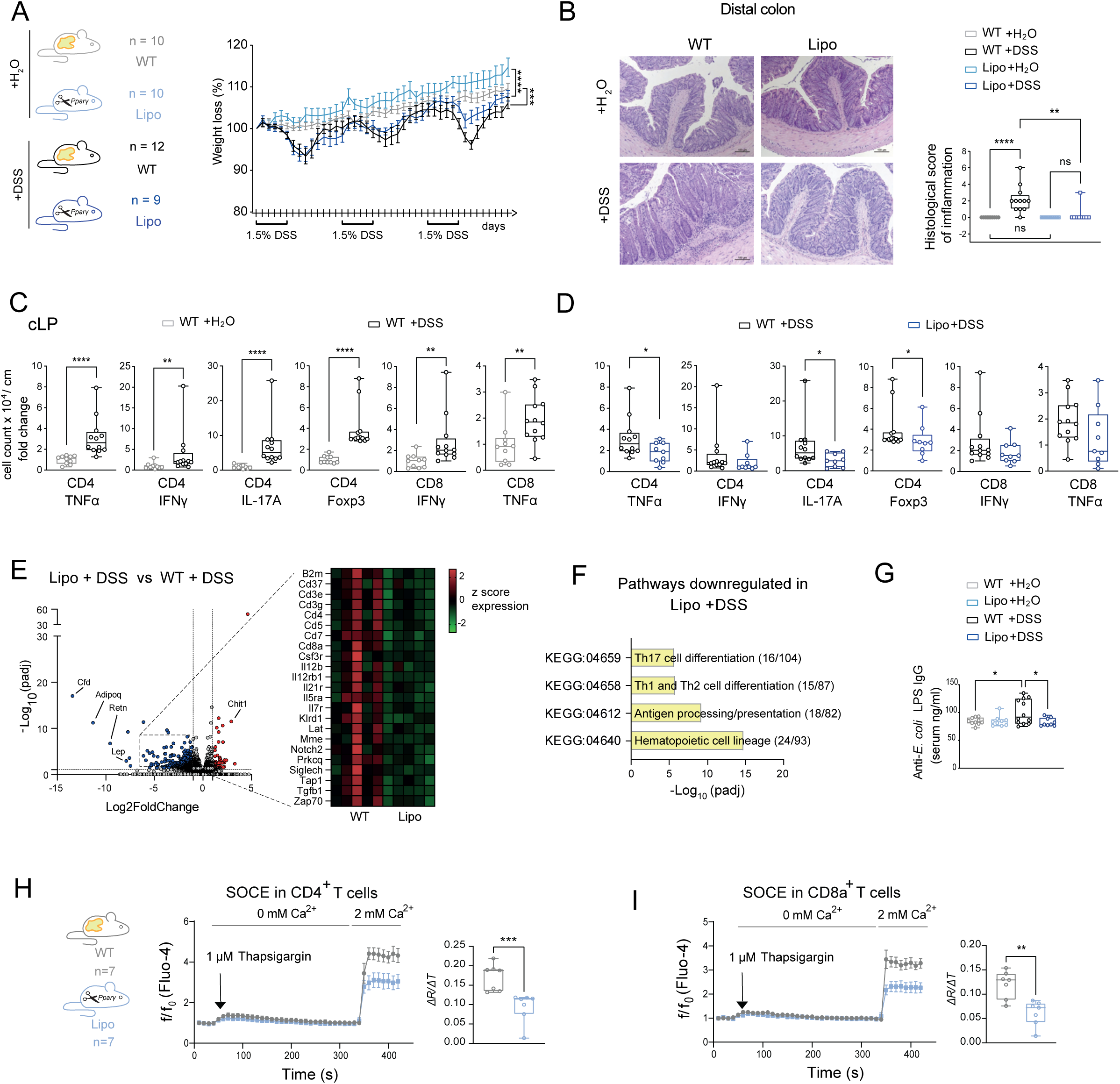
Lipodystrophic mice are protected from DSS-induced colitis. (**A**) Experimental groups for the induction of chronic DSS colitis and relative weight loss. Statistical differences were calculated by a one-way ANOVA test corrected for multiple comparisons according to Šídák’s method. Lines are projections of mean values. (**B**) Histologic score of colitis with representative H&E staining of distal colon sections (WT + H_2_O: n=10; WT+ DSS: n=12; Lipo +H_2_O: n=10; Lipo + DSS: n=9). Statistical differences were calculated using the Kruskal-Wallis test corrected for multiple comparisons with Dunn’s method. Data was pooled from two independent experiments. (**C**) Relative number of cells per cm of colon tissue in WT + DSS (n=12) versus WT + H_2_O: n=10, and (**D**) Lipo + DSS (n=9) vs WT + DSS: n=12 mice as assessed by flow cytometry. Statistical differences were calculated using the Mann-Whitney test. Data was pooled from two independent experiments. (**E**) Volcano plot showing differently regulated genes between Lipo and WT mice (n=6 per group) and heat map showing downregulated genes relating to inflammation in DSS-treated Lipo mice compared to DSS-treated WT controls. (**F**) KEGG pathway analysis and representative genes significantly downregulated (Padj <0.05 and Log2 fold-change <-1) in Lipo *+* DSS (n=5) compared to WT + DSS (n=5) mice. (**G**) Serum levels of anti-*E. coli* LPS IgG (WT + H_2_O: n=10; WT+DSS: n=12; Lipo +H_2_O: n=10; Lipo +DSS: n=9). Statistical differences were calculated by one-way ANOVA corrected for multiple comparisons with Šídák’s test. Boxes range from the 25th to the 75th percentile. Whisker plots show the minimum (smallest) and maximum (largest) values. The line in the box indicates the median. (SEM). Calcium influx upon treatment with Thapsigargin in splenic CD4^+^ (**H**), and CD8^+^ T cells (**I**) obtained from Lipo mice or wildtype littermates as assessed by flow cytometry. Data was pooled from two independent experiments (n=7 per group). P-value: ****P < 0.0001, ***P < 0.001, **P < 0.01, *P < 0.05.

Flow cytometric analyses of colonic immune cells revealed that DSS treatment effectively induced colitis in WT mice, which harbored significantly elevated numbers of CD4^+^ Tregs and proinflammatory CD4^+^/CD8^+^ T cell subsets compared to WT littermates receiving only water (**Figure 4C**). Notably, lipodystrophic mice treated with DSS were characterized by a significant reduction in colonic CD4^+^ T cells, producing TNFα or IL-17A, and a decline in Tregs (**Figure 4D**) in comparison to WT mice treated with DSS. Additionally, there was a notable decrease in intestinal CD8^+^ T cells in lipodystrophic mice, which further supports the notion that fat-derived signals play a pivotal role in the differentiation and function of proinflammatory T cells during intestinal inflammation. Consistently, bulk RNA sequencing (RNA-seq) of colonic tissue substantiated that DSS-treated lipodystrophic mice feature a significantly decreased expression of gene sets linked to T cell differentiation and antigen presentation when compared to DSS-treated WT littermates (**Figure 4E-F**). In addition, further gene set enrichment analyses (GSEA) not only revealed a significant decline in the expression of genes associated with inflammation but also showed a reduced epithelial-mesenchymal transition signature in DSS-treated lipodystrophic mice (**Suppl. Figure 4B**), indicating a preserved intestinal epithelial polarity and adherence in the absence of fat. Notably, we also observed that DSS-treated lipodystrophic mice had significantly less bacterial translocation through the intestinal barrier as fat-deficient mice harbored lower serum levels of anti-*E. coli* LPS IgG compared to WT mice receiving DSS (**Figure 4G**).

To better understand why T cells from lipodystrophic mice exhibit reduced cytokine production in the absence of adipose tissue, we conducted a comparative analysis of the ability of CD4^+^ and CD8^+^ T cells to flux Ca^2+^ through store-operated calcium entry (SOCE) between T cells obtained from lipodystrophic mice or WT littermates. SOCE, mediated by ORAI and STIM molecules, is a key signaling pathway controlling proinflammatory T cell function in murine and human IBD downstream of T cell receptor (TCR) activation^16^. As shown in **Figure 4H-I**, we observed that SOCE was significantly reduced in splenic CD4^+^ and CD8^+^ T cells derived from lipodystrophic mice when compared to T cells from WT mice. This suggests that T cells from lipodystrophic mice have a defect in the homeostasis of SOCE, which may contribute to impaired T cell function ultimately resulting in a decreased production of pro-inflammatory cytokines and a reduction in intestinal inflammation, as SOCE signaling is required for the production of IFNγ, IL-17A and TNFα in both CD4^+^ and CD8^+^ T cells and genetic or pharmacologic ablation of SOCE results in a reduced differentiation of Th1, Th17 and Treg cells^16^.

### Transplantation of allogeneic fat tissue partially rescues intestinal inflammation in lipodystrophic mice via a leptin-dependent induction of pro-inflammatory T cells

To further support our observation that adipose tissue is required for proper T cell homeostasis, we next examined how the reintroduction of adipose tissue would affect the T cell compartment of lipodystrophic mice under steady-state and inflammatory conditions. Using allogeneic adipose tissue transplantation models, we transplanted DSS-treated mice with either adipose tissue of WT donor mice or with adipose tissue of leptin-deficient *ob/ob* donor mice (**Figure 5A**, **Suppl. Figure 5A**). We observed that transplanted adipose tissue showed good engraftment with spontaneous vascularization, led to fat uptake in transplanted adipose tissue and partially restored basal leptin production in recipient lipodystrophic mice (**Figure 5A-C**; **Suppl. Figure 5B**). As expected, leptin production could thereby only be observed in recipient lipodystrophic mice receiving adipose tissue from WT donor mice but not in lipodystrophic mice transplanted with fat tissue from leptin-deficient *ob/ob* mice (**Figure 5C**). Remarkably, fat transplantation rescued the hepatic phenotype of lipodystrophic mice by reducing lipid load and liver weight (**Figure 5D-E**; **Suppl. Figure 5C-D**) underscoring the engraftment of functional adipose tissue after fat transplantation. Furthermore, western blot analyses of colonic tissue revealed that WT fat transplantation could partially reverse the overexpression of colonic TJ proteins observed in lipodystrophic mice under steady-state conditions (**Suppl. Figure 5E-F**), indicating that epithelial barrier functions decrease with increasing fat mass.

**Figure 5.**
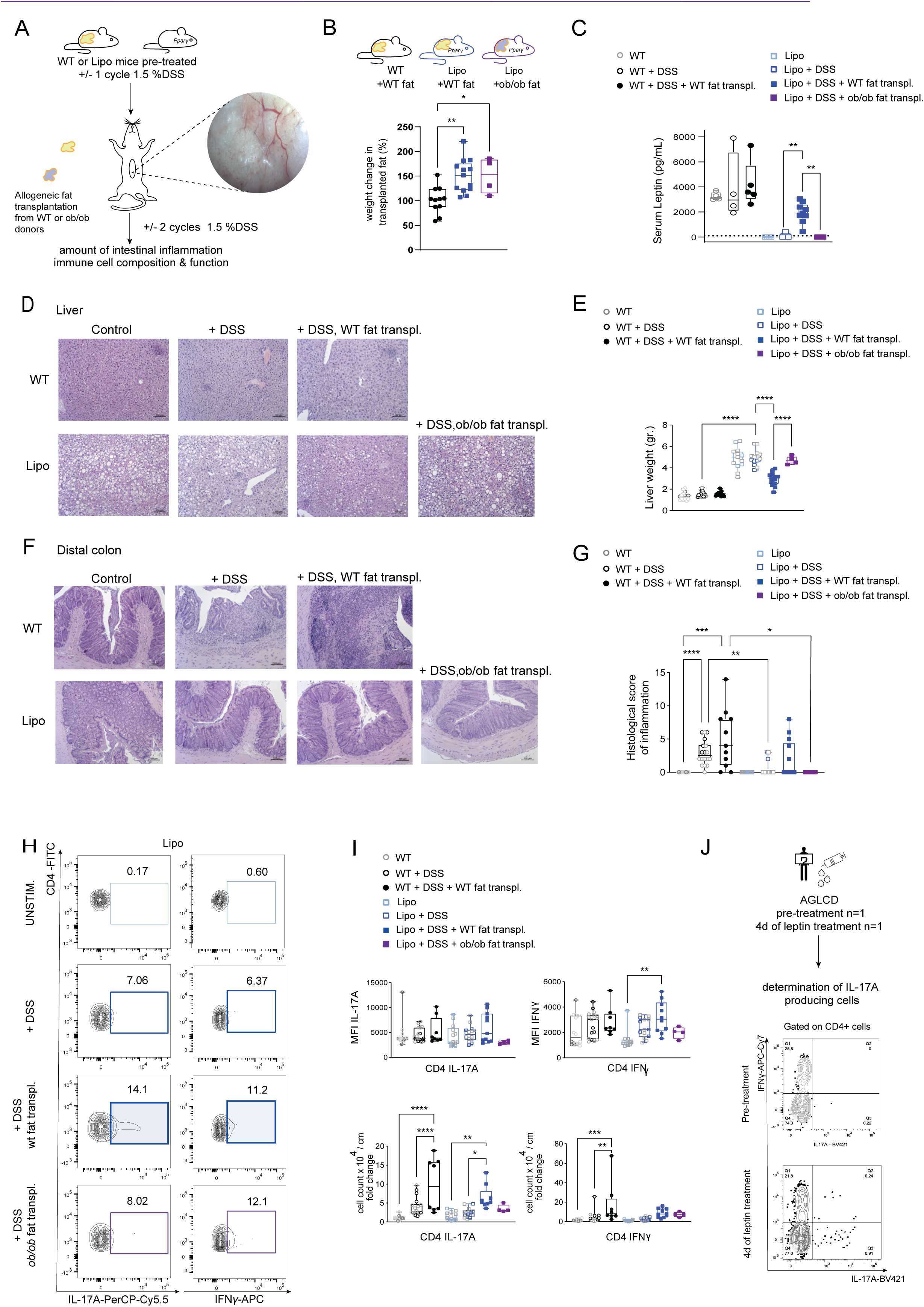
Allogeneic fat transplantation restores basal leptin levels, reverses steatohepatitis and promotes intestinal inflammation via the induction of intestinal proinflammatory T cells. (**A**) Experimental design: After receiving one cycle with 1.5% DSS, lipodystrophic (Lipo) or wild-type (WT) mice were transplanted with 500-600 mg adipose tissue obtained from WT or leptin-deficient *ob/ob* donor mice by performing mini-laparotomy. Seven days after transplantation, mice were subsequently challenged with 2 cycles of DSS treatment and sacrificed for organ harvesting. The image on the right shows vascularized transplanted fat tissue collected one month after surgery. (**B**) Bar graphs showing weight change of transplanted fat tissue relative to the initial weight of transplanted fat. (**C**) Serum levels of leptin = in transplanted versus non-transplanted animals (n=4-9). (**D**) Representative H&E staining of liver sections obtained from fat-transplanted versus non-transplanted animals. (**E**) Box-and-whisker plots showing the liver weight of fat-transplanted WT and Lipo mice (n=4-18). Data was pooled from 5 independent experiments. (**F**) Representative H&E staining of colon sections of DSS-treated animals after fat-transplantation in comparison to non-transplanted animals (n=4-18). (**G**) Box-and-whisker plots summarizing the histologic inflammation score of fat-transplanted and non-transplanted animals. Data shown is pooled from two independent transplantation experiments (bold symbols) and additional control data points derived from non-transplantation DSS experiments (light grey) shown in Figure 3 (n=4-18). (**H**) Representative FACS plots displaying the production of IFNγ and IL-17A in colonic CD4^+^ T cells from transplanted and non-transplanted animals. (**I**) Box- and-whisker plots summarizing the mean fluorescence intensity (MFI) of IFNγ and IL-17A in colonic CD4^+^ T cells as well as the absolute numbers of or IFNγ and IL-17A producing colonic CD4^+^ T cells in transplanted and non-transplanted animals normalized to WT mice. Data was pooled from 5 independent experiments (n=4-18). (**J**) Experimental setup and FACS plots showing the abundance of IFNγ and IL-17A-producing CD4^+^ T cells in the peripheral blood of the AGLCD patient before and 4 days after daily substitution with recombinant leptin. Statistical differences were calculated by one-way ANOVA test corrected for multiple comparisons according to Šídák’s method. Each point represents a single mouse, and the error bars represent the standard error mean (SEM) of one experiment. If significant, the p-value is given as: ****P < 0.0001, ***P < 0.001, **P < 0.01, *P < 0.05.

As shown in **Figure 5F-G**, the transplantation of WT adipose tissue resulted in comparable levels of histologic inflammation and weight loss (**Suppl. Figure 6A-B**) in WT and lipodystrophic mice treated with DSS. However, lipodystrophic mice that did not undergo fat transplantation or received fat from leptin-deficient *ob/ob* mice showed significantly reduced inflammation or no inflammation upon DSS treatment when compared to DSS-treated WT mice that had undergone transplantation with WT adipose tissue (**Figure 5F-G**). By analyzing RNA bulk sequencing data of colon tissue, we observed that the transplantation of WT fat tissue led to an enrichment of a pro-inflammatory Th1/Th2 gene signature in DSS-treated lipodystrophic mice when compared to DSS-treated lipodystrophic mice not receiving adipose tissue (**Suppl. Figure 6C-D**). Interestingly, we furthermore observed that the transplantation of WT fat in lipodystrophic mice caused an enrichment of a Th17 mRNA signature when compared to lipodystrophic mice receiving fat tissue from *ob/ob* mice (**Suppl. Figure 6E/F**), underscoring that fat-secreted leptin plays an important role in the differentiation of proinflammatory T cells. Likewise, we could detect a significantly higher number of colonic lamina propria Th17 cells in lipodystrophic mice receiving WT fat (**Figures 5H-I**), and we observed that the transplantation of WT fat significantly increased the production of INFγ in T cells of lipodystrophic mice (**Figures 5I**). Taken together, these findings confirm previous observations that fat-secreted leptin controls the differentiation of intestinal T cells in mice^13^. To confirm that leptin is not only required for T cell homeostasis in mice but also involved in the functional regulation of proinflammatory CD4^+^ T cells in humans, we analyzed PBMCs from our AGLCD patient, obtained before treatment with recombinant leptin and following 4 days of daily treatments with recombinant leptin. Consistent with our observations in mice, we found that recombinant leptin treatment increased the abundance and the expression of IL-17A-producing CD4^+^ T cells in the AGLCD patient *in vivo* (**Figure 5J**). Taken together, these findings demonstrate that fat-derived signals such as leptin are required for the differentiation and function of pro-inflammatory T cells in both mice and humans and that lipodystrophy strongly protects against intestinal inflammation.

### Identification of a highly restricted T cell receptor repertoire in the AGLCD patient

Given the high frequency of CD8^+^ T cells that we had observed in the periphery as well as at the site of inflammation in the AGLCD patient despite the absence of fat-derived signals, we postulated that the observed expansion of T cells was primarily T cell-intrinsic rather than being caused by missing-fat derived signals. We thus decided to characterize the clonotypes of the AGLCD patient via single-cell sequencing. Single-cell TCR sequencing of sorted peripheral CD3^+^ T cells revealed an over-representation of oligoclonal, hyperexpanded T cells in the AGLCD patient (**Figure 6A-B**) in comparison to T cells from a healthy donor. Mapping the top 10 expanded clonotypes onto the scRNA-seq data showed that this phenomenon was particularly pronounced in effector memory CD4^+^ and CD8^+^ T cells and MAIT cells (**Figure 6C**). However, none of the identified sequences overlapped with annotated TCR sequences present in pre-existing IBD cohorts (whole blood samples from 108 CD, 36 patients with ulcerative colitis (UC), 99 HD or intestinal surgical specimens from 11 CD, 13 UC, and 13 non-inflamed controls)^29^ or in the VDJ database^30^. To understand if these clonally restricted T cells persist over time and contribute to intestinal inflammation in the AGLCD patient, we sorted CD4^+^ and CD8^+^ T cells from both the blood and inflamed ileal tissue at two different time points six months apart. We then analyzed their TCR sequences using bulk sequencing. By comparing TCR sequences from single-cell TCR data with those from bulk sequencing, we found overlapping TCR sequences in both blood and inflamed tissue, particularly in the central- and effector memory CD4^+^ and CD8^+^ T cell compartment, as well as in MAIT cells (**Figure 6D-E**). Additional differential gene expression analyses showed that these T cells with overlapping TCRs between blood and intestine featured an inflammatory phenotype, with high expression levels of IFNγ and TNFα as well as a significant enrichment of gene signatures related to TNFα, IFNγ, IL-2, IL-6, and mTORC1 signaling (**Figure 6F-H**). Remarkably, these clonally restricted cells were furthermore characterized by a high expression of genes related to lipotoxic stress, such as APOO, and a gene set related to cholesterol homeostasis (**Figure 6G-H**), highlighting that cells are exposed to a high burden of lipids in the absence of adipose tissue. Taken together these observations indicate that the AGLCD patient chronically accumulates clonally stable inflammatory T cells both at the site of inflammation and in the periphery.

**Figure 6.**
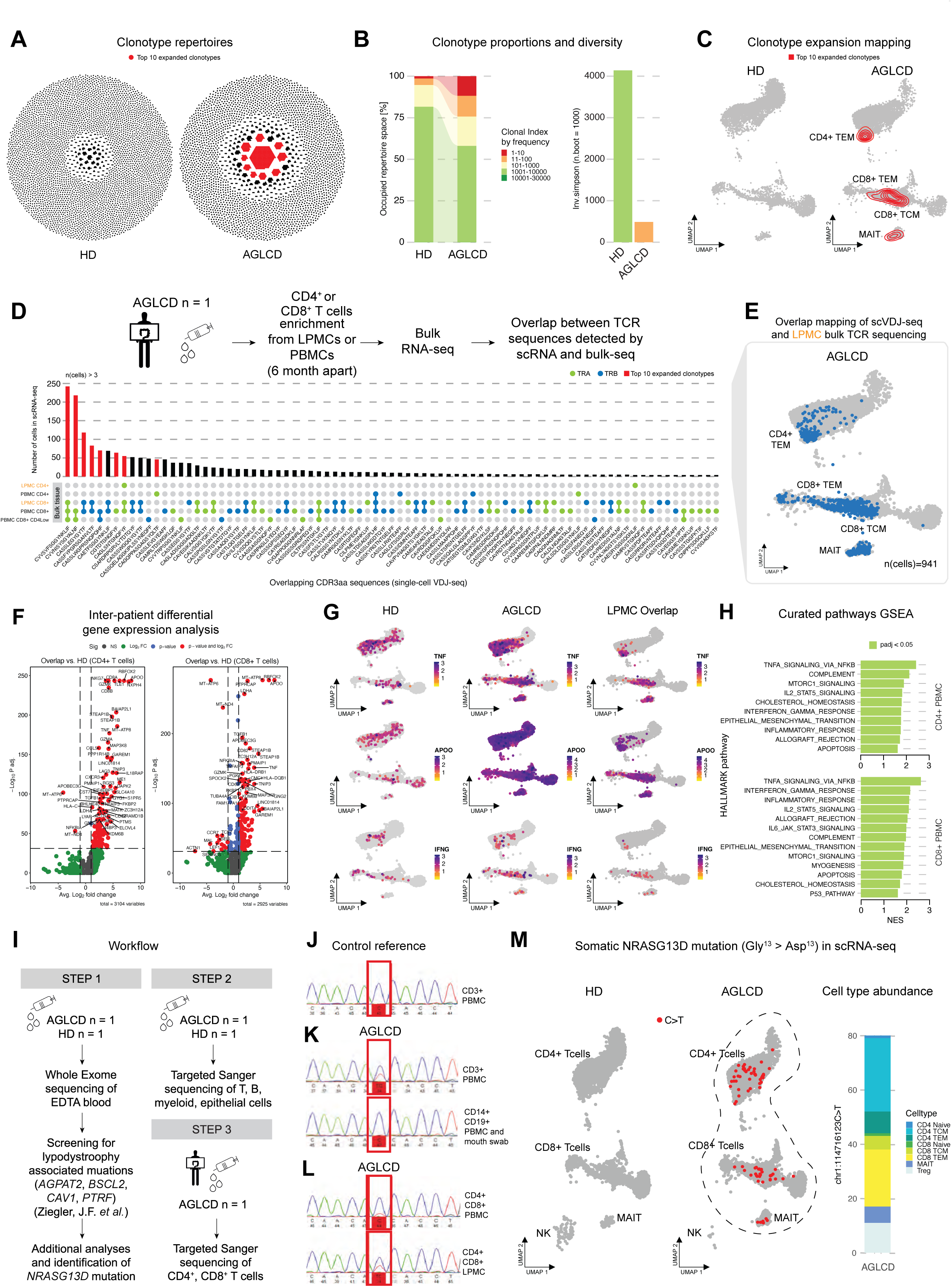
Detection of a somatic *NRASG13D* mutation in clonally-restricted T cells of the AGLCD patient. **(A)** Honeycomb plots derived from scTCR sequencing of 10,000 T cells obtained from the AGLCD patient or 1 healthy donor (left) showing the clonal distribution of T cells. (**B**) Stacked Bar plot depicting clonal proportions of the occupied repertoire space in the AGLCD patient compared to a healthy donor (HD) and Bar plot visualizing the inverted Simson Index for T cell clonality of the AGLCD patient in comparison to the HD (results obtained from 1000 bootstrap resamples). (**C**) UMAPs displaying density mapping of top 10 expanded clonotypes in T cell subsets of the AGLCD patient. (**D**) Overlap analysis of T cell CDR3aa sequences from scTCR sequencing of PBMCs and bulk TCR sequencing of sorted intestinal and PBMC-derived T cells from the AGLCD patient at two time points (6 months apart). The bar plot shows the number of cells with matching CDR3aa sequences in the scRNA-seq data linking all 3 data sets. UMAP maps a subset of cells from PBMC-derived scRNA-seq data containing CDR3aa chains overlapping with the site of inflammation (LPMCs). (**E**) Volcano plots showing differentially expressed genes in CD4^+^ and CD8^+^ T cells with TCR-overlap between blood and inflamed lamina propria of the AGLCD patients compared to T cells of an HD. (**F**) UMAPs showing non-zero TNF, IFNG and APOO mRNA expression in the HD (left), all cells of the AGLCD patient (center) and clonally overlapping T cell subsets (right). (**G**) GSEA pathway analyses in clonally overlapping CD4^+^ and CD8^+^ T cell subsets of the AGLCD patient. (**H**) Experimental setup of DNA sequencing experiments. (**I-K**) Targeted Sanger sequencing of the *NRAS* locus in buccal epithelial cells, CD3^+^ T cells, CD19^+^ B cells and CD14^+^ myeloid cells sorted from peripheral blood or CD4^+^ and CD8^+^ T cells isolated ileal lamina propria (LP) of the AGLCD patient. (**L**) UMAPs obtained from scRNA sequencing show the abundance of the *NRASG13D* mutation in T cell subtypes (**J**).

### T cells of the AGLCD patient carry a somatic de novo NRASG13D mutation

Prior research has indicated that somatic *de novo* mutations in T cells may play a role in the survival and evolution of clonally restricted T cells in patients with rheumatoid arthritis, multiple sclerosis, and non-malignant hematologic or immunologic disorders, including aplastic anemia, myelodysplastic syndrome, and autoimmune thrombocytopenia^31, 32, 33^. To understand whether the observed long-term persistence of clonally expanded T could be supported by the occurrence of somatic mutations in the AGLCD patient, we next examined the mutational landscape of lymphocytes in the AGLCD patient. Using whole exome sequencing (WES) generated from EDTA blood of the AGLCD patient and his relatives^11^, we had previously excluded germline mutations commonly associated with inherited forms of generalized lipodystrophies in genes regulating lipolysis and adipocyte homeostasis (*AGPAT2*, *BSCL2*, *CAV1, PTRF*)^34, 35, 36, 37^. However, by looking beyond these genes and further re-analyzing this existing WES data set^11^, we were able to detect a *de novo* mutation causing a non-conservative aspartic acid to glycine substitution at codon 13 in the *NRAS* gene (*NRASG13D*) (**Figure 6I**, STEP1). The *NRAS* gene encodes the NRAS protein, an essential regulator of the hematopoietic system development^38^.

For example, NRAS was found to play a crucial role during the development and function of CD8^+^ T cells in mice as NRAS-deficient CD8^+^ T cells show impaired thymic development and functional impairments in controlling infections with influenza virus^39^. Previous studies have furthermore identified mutations that lead to the constitutive activation of NRAS as a potential risk factor for hematopoietic malignancies in patients and transgenic mouse models^40^. In particular, the *NRASG13D* mutation has previously been implicated in the pathogenesis of autoimmune-lymphoproliferative syndromes (ALPS), by turning T cells and lymphocytes more resilient to growth factor deprivation and by causing defects in the induction of apoptosis, ultimately resulting in a systemic non-malignant-lymphocyte-expansion^15^. Since the *NRASG13D* mutation was detected in less than 10% of all reads by WES performed on whole blood cells (data not shown), we hypothesized that only a small fraction of PBMCs might carry the *NRASG13D* mutation. To test this, we used targeted Sanger sequencing to separately analyze peripheral CD3^+^ T cells, CD19^+^ B cells, CD14^+^ monocytes, and buccal epithelial cells isolated from the AGLCD patient or a healthy donor (HD) (**Figure 6I**, STEP2). As shown in **Figure 6J-L**, from all the cell types investigated, only CD3^+^ T cells harbored the *NRASG13D* mutation. Further analysis of sorted CD4^+^ and CD8^+^ T cells isolated from either blood or intestinal biopsies of the AGLCD patient (**Figure 6I**, STEP3) could confirm the presence of the *NRASG13D* point mutation in T cells in the peripheral blood compartment (**Figure 6K**) as well as in FACS-sorted T cells at the site of inflammation in the intestine (**Figure 6K**). Remarkably, we were also able to detect RNA reads spanning the *NRASG13D* mutation in our scRNA-seq data in CD4^+^ and CD8^+^ T as well as in MAIT cells but not in NK and B cells, supporting the notion that the AGLCD patient carries a *de novo NRASG13D* mutation exclusively in the T cell compartment (**Figure 6M**). In light of the growing evidence for the role of somatic mutations in the evolution and maintenance of clonally restricted T cells in non-malignant T cells, our data collectively indicate that the occurrence of the *NRASG13D* mutation might facilitate the survival of autoreactive T cells, thereby advancing the selection and survival of clonally restricted inflammatory T cells in the AGLCD patient ultimately contributing to persistent intestinal and systemic inflammation, which can then be further aggravated by exogenous substitution with recombinant leptin.

## Discussion

In summary, we here describe a T cell-intrinsic lymphoproliferation characterized by a heterozygous *de novo* mutation of *NRAS* as a potential driver of systemic and intestinal inflammation in AGL. A T cell-driven cause of AGL is thereby supported by observations in cancer patients who develop AGL after treatment with the immune checkpoint blocking antibodies pembrolizumab and nivolumab, resulting in adipose tissue infiltration by CD8^+^ T cells, progressive generalized lipodystrophy, severe insulin resistance, and general autoimmunity^4, 5, 6^.

Using mouse models of lipodystrophy, we were able to exclude that the observed lymphoproliferation in AGLCD is a secondary consequence of absent fat-derived adipokine signaling, as we did not observe a spontaneous development of lymphoproliferation in lipodystrophic mice. In contrast, we demonstrated that lipodystrophy protects mice from chronic DSS-induced intestinal inflammation by reducing the differentiation of pro-inflammatory T cell subsets, especially Th1 and Th17 cells, and by improving intestinal barrier properties. Vice versa, the transplantation of allogenic fat tissue resulted in a decreased expression of tight junction proteins such as Cldn3 and angulin-1/LSR proteins and promoted the differentiation of pro-inflammatory T cells in a leptin-dependent manner, ultimately exacerbating intestinal inflammation. These results align with our prior observations, where we noticed a worsening of intestinal inflammation in the AGLCD patient after treatment with recombinant leptin, which eventually necessitated an aggressive triple immune suppression using the TNF-blocker adalimumab, tacrolimus, and MMF for the treatment of autoimmunity in the AGLCD patient^11^. In summary, our data suggest that leptin may act as a pro-inflammatory adipokine in AGLCD, further aggravating both systemic and intestinal auto-inflammation which is primarily driven by a T cell-intrinsic lymphoproliferation of inflammatory CD4^+^ and CD8^+^ T cells.

Due to the previously reported role of *NRASG13D* in the promotion of T cell lymphoproliferation in patients with ALPS^15^ and the highly restricted TCR repertoire in T cells of the AGLCD patient, it seems very likely that the presence of *NRASG13D* facilitates the survival and/or clonal evolution of inflammatory T cells thereby promoting autoimmunity and systemic inflammation. In this study, we were able to detect the stable occurrence of the same clonally restricted effector memory CD4^+^ and CD8^+^ T cells both in the periphery as well as at the site of inflammation in the gut of the AGLCD patient over time. We could also observe that these clonally restricted cells carry the *NRASG13D* mutation, express high levels of inflammatory markers such as IFNγ and TNFα, and display a significant enrichment of gene-sets related to mTORC1 signaling, which is a crucial pathway for maintaining immune receptor signaling, effector functions and metabolic homeostasis of T cells^41^. Taken together, these findings suggest that the occurrence of somatic mutations in clonally restricted cells might contribute to the persistence of clonally restricted inflammatory T cell subsets in AGL over time. Likewise, Lundgren and colleagues recently identified an *NRASG13D* mutation as a potential driver of clonal T cell expansion in a patient with aplastic anemia^31^, which is supported by observations from Garcia and colleagues, who have demonstrated that naturally occurring somatic mutations can enhance the function and survival of CAR T cells *in vitro* and *in vivo*^42^. Moreover, clonally restricted T cells were found to harbor somatic mutations in up to 20% of patients with other autoimmune diseases such as rheumatoid arthritis and multiple sclerosis^32, 33^. However, it remains speculative which antigens are recognized by the hyperexpanded T cells found in the AGLCD patient, and future studies will be required to delineate how somatic mutations such as the *NRASG13D* mutation contribute to the selection of antigen-specific inflammatory cells under sub-optimal conditions such as hypoxia and inflammation, both in AGL and in autoimmune diseases in general.

A T cell-driven mechanism in AGL is also supported by Fischer-Posovszky and colleagues, who previously reported that T cells induced apoptosis of adipocytes in a FASL-dependent manner in a young patient with autoimmune-associated AGL^43^. Despite this observation, the specific antigens responsible for T cell activation in the adipose tissue of AGL patients are unknown and it remains speculative if the autoreactive T cells found in the periphery and gut of the AGLCD patient can also contribute to adipose tissue destruction, as we were unable to isolate fat-residing T cells from the AGLCD patient due to the patient’s complete absence of adipose tissue. However, given the established role of T cells in autoimmune AGL, it is in our opinion still plausible to hypothesize that the observed T cell proliferation may drive both autoimmune lipodystrophy and intestinal inflammation in the AGLCD patient. Similarly, concomitant local inflammation of adipose tissue such as panniculitis and erythema nodosum^44^ can also observed in Crohn’s disease patients with extraintestinal manifestations, suggesting that the complete loss of fat tissue in the AGLCD patient might also be viewed as a very extreme form of an extraintestinal manifestations of Crohn’s disease.

Additionally, our data caution that the *de novo* mutation in *NRAS* may facilitate the transformation of lymphoproliferative T cells into malignant lymphoma, given the high incidence of T cell lymphoma observed in patients with AGL^3^. This is particularly important because mutations in *NRAS* are associated with therapy resistance and unfavorable clinical outcomes in patients with cutaneous T cell lymphoma^45^. One limitation of this study stems from the prolonged disease course of our AGLCD patient and the intensive immunosuppressive therapy with tacrolimus, MMF, and TNF blockers. As a result, it remains speculative whether the somatic *NRASG13D* mutation is a consequence of the harsh immunosuppressive therapy, the patient’s severe inflammation or a fundamental driver in the development of AGLCD. Another factor that should be considered as a potential contributor to genetic instability and the subsequent formation of somatic mosaicism is the high lipid burden and the consecutive lipotoxic challenge that lymphocytes experience during AGL due to systemically elevated triglyceride levels. We could previously detect in the AGLCD patient that in the absence of adipose tissue monocytes, NK and CD8^+^ T cells store high amounts of lipid droplets^11^. By using scRNA sequencing, we were able to detect a significant upregulation of genes that have been associated with lipotoxicity including *APOO*, which has been previously described to act as an important mediator of lipotoxicity causing increased production of reactive oxygen species as well as disturbances in mitochondrial homeostasis^46^. It is thus conceivable that the elevated levels of circulating lipids observed in AGL may contribute to genetic instability, thereby not only supporting the formation of somatic mutations in inflammatory non-malignant T cells as evidenced in the current study but also increasing the overall probability of genetic alterations, ultimately leading to an increased risk for lymphoma development in AGL.

Despite recent advances in sequencing technologies, somatic mutations may often be missed by standard bulk sequencing approaches, as mutated cell lines often represent only a small fraction of cells in tissue conglomerates or whole blood samples. We believe that it will be critical to thoroughly consider single-cell analyses to delineate the intra-individual genetic heterogeneity driving complex diseases such as AGL and to develop individualized therapies based on these analyses. Whether somatic *de novo* mutations may be a specific feature of AGLCD or whether similar *de novo* mutations may underlie the development of inflammatory bowel disease in general, it is currently unknown and needs to be investigated in future studies.

We believe, that our data are not only crucial in the context of AGL as they show that autoimmunity in AGL is likely to arise from T cell-intrinsic events such as somatic mosaicism and clonal evolution of inflammatory cells, but they also may help to better delineate the so far incompletely understood function of adipose tissue in the pathogenesis of IBD. Although findings from cross-sectional and retrospective cohort studies correlating the effect of adipose tissue with the development of intestinal inflammation have indicated that obesity is associated with a higher risk of developing Crohn’s disease, the underlying mechanisms of this correlation remain unsolved^9^. Our data support the pro-inflammatory role of adipose tissue and its secreted factors in intestinal inflammation while confirming that adipose tissue-derived signals, and especially leptin, play an important role in the regulation of immune cell function by controlling the differentiation and function of inflammatory T cells. This suggests that the removal or reduction of adipose tissue with subsequent reduction in adipokine levels may improve intestinal barrier function and reduce the generation of proinflammatory T cells such as Th17 and Th1 in patients with relapsing IBD. Overall, this study depicts a complex picture of the interplay between adipose tissue and immune cell homeostasis and argue that a tight regulation of fat-derived signals is required to prevent excessive autoimmune responses in IBD. In addition, our findings advocate that somatic mutations in T cells might contribute to the development of complex autoimmune phenotypes and that somatic mutations should be considered as potential drug targets in personalized treatment approaches of complex IBD with severe extraintestinal manifestation in the future.

## Data Availability

Patient data supporting the human findings of this study are available from the corresponding authors upon reasonable request. scRNA-seq and mouse bulk seq data for this project will be uploaded to the NCBI database upon acceptance of this paper.

## Acknowledgment

This work was funded by the German Research Foundation (We 5303/3-2 to CW, SFB-TRR 241 (project-ID 375876048) B01 to BS and CW, A09 to AS and CW, A03 to CB and Z02 to BS, CRU 5023 (project-ID: 50474582), CRC 1449-B04 (project-ID: 431232613); CRC 1340-B06 (project-ID 372486779), SI749/14-1 (project-ID: 418055832) all to BS. CW and JZ received funding from the Clinician Scientist Program of the Berlin Institute of Health. CW received funding by the Fritz-Thyssen Foundation (10.19.2.028MN). *Prarg^fl/fl^* mice were a kind gift of Prof. Dr. Ulrich Kintscher, Charité, Berlin.

## Materials and Methods

### Ethics and study approval

Written informed consent was obtained from all healthy volunteers and patients as approved by the institutional review board of Charité - Universitätsmedizin Berlin. Cohort characteristics are summarized in **Suppl. Table 1**.

### Isolation of peripheral blood mononuclear cells

Heparinized blood was collected at various time points from the AGLCD patients and healthy volunteers. First, peripheral blood mononuclear cells (PBMCs) were isolated by density gradient centrifugation using SepMate-50™ tubes (#85450, Stemcell Technologies, Vancouver, Canada) according to the manufacturer’s instructions. The cells were then washed twice in a complete medium. Finally, isolated PBMCs were either snap-frozen in FBS (Sigma-Aldrich) supplemented with 10% dimethyl sulfoxide (DMSO) (#D8418, Sigma-Aldrich) and stored in liquid nitrogen or used directly for *in vitro* assays.

### Isolation of lamina propria mononuclear cells

Surgical specimens of the terminal ileum were obtained from CD and the AGLCD patient who underwent colectomy. In contrast, non-inflamed specimens of patients with colorectal cancer served as controls. As previously described^16^, ileal mucosa was dissected, washed in 1 mM 1,4 dithiothreitol (DTT) (#6908.4, Carl Roth), and dissolved in Hanks’ balanced salt solution (HBSS) without Ca^2+^ and Mg^2+^ (#14175095, Thermofisher Scientific). Lamina propria tissue was cleaned of any left submucosa, cut into pieces, and washed three times in 1 mM EDTA (#E6511, Sigma-Aldrich) for 15 min at 37 °C in shaking condition at 250 rpm. The EDTA was then washed out with HBSS without Ca^2+^ and Mg^2+^ (Thermofisher Scientific), and the tissue was incubated in a digestion medium supplemented with 0.15 mg/mL collagenase A (Roche) for 16 hours at 37 °C in shaking condition at 200 rpm (**Suppl. Table 2**). On the following day, the supernatant was filtered through a 100 µm cell strainer (#08-771-19, Thermofisher Scientific) and washed three times in HBSS without Ca^2+^ and Mg^2+^. Cells were separated by centrifugation on a Percoll gradient (#17089101, GE Healthcare, Chicago, Illinois) at 300 g for 30 min at 4 °C. Lymphocytes and monocytes were isolated from the 40-60% Percoll (GE Healthcare) interface, washed twice, and used for downstream assays.

### Mass cytometry staining and acquisition

LPMCs isolated from ileal resections were stimulated *ex vivo* for 4 h in activation medium (**Suppl. Table 2**). In addition, 10 µg/mL brefeldin A (#B6542, Sigma-Aldrich) was added to all samples for 2 h, while 25 KU benzonase nuclease (#E1014, Sigma-Aldrich) was added (1:10,000) 15 min before harvesting. LPMCs were then fixed in Smart Tube buffer (#PROT-1, SMART TUBE, Las Vegas, Nevada) supplemented with 20% BSA (#9048-46-8, Sigma-Aldrich) and subsequently stored at −80 °C until antibody staining and acquisition. On the day of acquisition, single-cell suspensions were thawed, and to pool multiple samples in a single batch/acquisition run (max 40 samples per batch/run) to avoid introducing batch effects, LPMCs were barcoded for 30 min at room temperature (RT) using the Cell-ID 20-plex Pd Barcoding Kit (#201060, Fluidigm) ± CD45-89Y staining. Individual samples were washed twice with cell staining buffer (#201068, Fluidigm) and pooled prior to staining. Anti-human antibodies were either purchased pre-conjugated to metal isotopes or conjugated in-house using the MaxPar X8 conjugation kit (#201300, Fluidigm). A complete list of antibodies used for mass cytometry staining is provided in **Suppl. Table 3**. Cells were incubated for surface staining for 30 min at 4 °C, washed twice with cell staining buffer, and incubated in fixation/permeabilization buffer (#88-8823-88, eBioscience) for 60 min at 4 °C. After two washes with permeabilization buffer (eBioscience), cells were stained with an antibody cocktail against intracellular molecules for 1 hour at 4°C. Cells were then washed twice with permeabilization buffer and incubated overnight in 2% methanol-free formaldehyde solution (#28908, Thermofisher Scientific) for 1 hour at room temperature. LPMCs were then washed and resuspended in 1 mL of iridium intercalator solution (#201192A, Fluidigm) for 1 hour at RT. Finally, cells were washed twice with cell staining buffer and then twice with double-distilled water (ddH_2_O) for time-of-flight (CyTOF) mass cytometry measurements. As described previously^47, 48^, cells were acquired using a CyTOF2 upgraded to Helios specifications (Fluidigm), at the BIH Flow & Mass Cytometry Core Facility, Berlin, Germany.

### Analysis of mass cytometry data

The Cytobank software package (www.cytobank.org) was used for the initial manual gating of single cells and the de-barcoding of pooled samples. FCS files of de-barcoded single cells were exported and further compensated for signal spillover using the R software (3.6.0, Bioconductor 9) (www.R-project.org) and by applying the CATALYST package and arcsinh transformation (scale factor 5) prior to data analysis^49^. Compensated files were then gated for CD45^+^ cells (**Suppl. Figure 1**), and t-distributed stochastic neighbor embedding (t-SNE) maps were generated according to the expression of lineage and/or functional markers. t-SNE maps were generated according to the expression of linage markers for T (CD4, CD8, CD45RO, CD45RA, CCR7, IL-7R, CD27, CD25, PD-1, perforin), B (CD38, CD19), NK (CD56) and myeloid cells (CD11b, CD11c) (**Figure 1C, D**). For all analyses, FCS files with t-SNE embedding as two additional parameters were exported from Cytobank. Clustering analysis was performed using the *FlowSOM/ConsensusClusterPlus* package in R (3.6.0, Bioconductor 9)^50^. Cell clusters of LPMCs were identified after visual inspection of t-SNE plots and functional interpretation of heat maps generated by *FlowSOM/ConsensusClusterPlus*. Statistical analysis of differential abundance (DA) clusters between disease groups was performed using a generalized linear mixed effects model (GLMM) available through the R package diffcyt (using all defaults with analysis_type = ”DA,” method_DA = ”diffcyt-DA-GLMM,” min-cells = 3). The false discovery rate (FDR) was adjusted to 10% using the Benjamini-Hochberg (BH) procedure for multiple hypothesis testing, as previously described^51^. Multiple t-test (Benjamini, Krieger, and Yekutieli two-stage linear step-up procedure, with Q = 1%) was used to analyze protein expression levels among disease groups.

### Whole exome sequencing

PBMCs were obtained as described, and CD3^+^, CD8^+^, CD4^+^ T cells, CD19^+^ B cells, and CD14^+^ myeloid cells were sorted using either with anti-allophycocyanin (APC) (#130-090-855) or anti-FITC MultiSort Kits (#130-058-701, Miltenyi Biotec) according to the manufacturer’s instructions or using the FACSJazz™ Cell Sorter (BD). A complete list of antibodies used for sorting can be found in **Suppl. Table 4**. DNA was extracted from PBMC subsets or buccal swaps using DNeasy Blood & Tissue Kits (#69504, Qiagen, North Rhine-Westphalia, Germany). Exome sequencing was performed by the Institute of Clinical Molecular Biology, Kiel, Germany, using the xGen Exome Research Panel v 1.0 and 2 × 75 bp paired-end, while the sequencing was carried out on an Illumina HiSeq 3000 (Illumina), as previously reported^11, 48^. Reads were mapped to the human reference genome assembly hg19 using BWA^52^. They were then sorted, converted to bam format, and indexed using SAMtools^52^. This was followed by PCR duplicate removal (http://picard.sourceforge.net) and local realignment around InDels followed together with base quality score recalibration using GATK according to their best practice recommendations, in accordance with variant calling and variant quality score recalibration. Finally, Alissa Interpret (Agilent) was used for Variant annotation and filtering.

### Single cell RNA sequencing

PBMCs were collected from the AGLCD patient and a healthy donor, as described above, and resuspended in 1X PBS (#10010023, Gibco) containing 2% FBS (Sigma-Aldrich) and 1 mM EDTA (#E6511, Sigma-Aldrich) at a density of 5 x 10^7^ cells/mL. CD3^+^ T cells were subsequentially isolated using the EasySep™ Human T Cell Isolation Kit (#17951, Stemcell Technologies) and transported to the Center for Molecular Biosciences (ZMB), Kiel, Germany. CD3^+^ single cell suspensions were loaded onto a Chromium Chip G (10x Genomics, Pleasanton, California) according to the manufacturer’s instructions for processing with the Chromium Next GEM Single Cell 5’ Library and Gel Bead Kit v1.1. 15,000 cells were loaded for each reaction. TCR single cell libraries were subsequently generated from the same cells using the Chromium Single Cell V(D)J Enrichment Kit, Human T Cell (10x Genomics). Libraries were sequenced on the Illumina NovaSeq 6000 machine (Illumina) using 2×100 bp for gene expression, aiming for 50,000 reads per cell and 2×150 bp and 5000 reads per cell for TCR libraries. Pre-processing of scRNA-seq data was performed with the Cell Ranger software (v7.1.0; 10x Genomics) using the human genome reference GRCh38 (v7.1.0) for mapping. Ambient RNA was removed using Cellbender and doublets filtered assuming a 7.5% doublet formation rate using the R package DoubletFinder (vbefore standard scRNA-seq preprocessing in the R package Seurat (v5.1.0; min.cells = 3, min.features = 800). Single-cell TCR distribution and clonal expansion were visualized using enclone (10x Genomics) and the R package scRepertoire (v2.0.4).

### Bulk TCR repertoire

PBMCs or LPMCs were collected as described above, and CD8^+^ or CD4^+^ T cells were sorted using the FACSJazz™ Cell Sorter (BD). A complete list of antibodies used for sorting can be found in **Suppl. Table 4**. Total RNA was isolated using the Rneasy Mini Kit (#74104, Qiagen). Subsequently, molecular barcoded TCR cDNA libraries were prepared by the ZMB Center of Kiel, Germany, as previously described^53^, with minor modifications for both TCRα and TCRβ chains. Briefly, cDNA synthesis was performed using SMARTScribe reverse transcriptase (Clontech, Mountain View, CA) with primers for the TCRα and TCRβ constant regions. A unique molecular identifier (UMI), and a 6-nucleotide sample barcode were introduced via template switching. cDNA synthesis was performed 42 °C for 60 min. cDNA was then treated with Uracil DNA-Glycosylase (UDG) (New England Biolabs, Ipswich, MA) and incubated for 30 min at 37 °C. Samples were then purified using the QIAquick PCR Purification Kit (Qiagen) and eluted in 50 or 100 µl of deionized water. Purified cDNA was amplified in two consecutive PCRs, 18 and 12 cycles, with purification after each PCR using MagSi-NGSprep Plus (MagnaMedics, Aachen, Germany). Illumina-compatible adapters and sample-specific barcodes were added during the second PCR. Library quality and concentration were measured using TapeStation D1000 (Agilent) and Qubit (Thermofisher Scientific). Five ng per library were pooled and sequenced on Illumina MiSeq v2 with a single index run at 2×150 bpm (Illumina). Custom sequencing primers were added to the Illumina primers. Bulk TCR data were aligned to the human TCR gene reference (repseqio.v4.0) in a quality-aware manner to correct for sequencing and PCR errors. Clonotypes were then assembled and grouped by their CDR3 sequences using the software MiXCR^54^ (v4.6.0; preset “rna-seq”). Clonotype tables containing clonotype counts, frequencies, CDR3 nucleotide, amino acid sequences, and V(D)J genes were generated and used for further analysis.

### Integration of single-cell and bulk TCR repertoire datasets

Single-cell alpha and beta TCRs were separated to emulate a pseudo-bulk repertoire and compared with the bulk TCR repertoire data from the ZMB Center in Kiel, Germany. Alpha diversity measures such as the proportion of clonotypes and the inverse Simpson index were calculated using the R software package scRepertoire (v2.0.4). The presence and frequency of clonotypes between bulk and single cell data was then compared within the respective CD4^+^ and CD8^+^ cell fractions of the AGLCD patient. In addition, TCRs were searched in the VDJdb database^30^ for antigen and HLA specificity. To check for possible TCR overlap in pre-existing patient cohorts^29^, we compared the clonotypes identified in the AGLCD patient with whole blood samples from 108 CD, 36 UC, 99 healthy, and surgical specimens from 11 CD (4x colon, 7x small bowel), 13 UC, 13 colorectal cancer (CRC), all from the colon.

### Mice ethics

While *Pparγ^fl/fl^-Adipoq-Cre* lipodystrophic mice were bred at the animal facility of the Charité, *ob/ob* mice were purchased from Charles River, Germany. All experiments were performed according to protocols approved by Berlin’s Regional Animal Experimentation Committee (LaGeSo), and to reduce animal burden, both female and male rodents were included in this study.

### DSS colitis

Lipodystrophic mice and control littermates, 9 to 13 weeks of age, were given water with or without 1.5% DSS (MP Biomedicals) for five days, followed by nine days of plain water, repeated for a total of three cycles. Mice were monitored daily for weight loss and consistency/presence of blood in the stool. Stool consistency was determined as follows: score 0, normal (solid pellet); 1, soft but in pellet shape; 2, loose stool but with some solidity; 3, loose stool with signs of liquid consistency; 4, watery diarrhea. Rectal bleeding was scored as follows: score 0, no sign of blood; score 1, no bleeding; 2, slight bleeding; 3, bloody diarrhea; 4, gross bleeding^9^. At the endpoint, organs were harvested and stored or processed directly for downstream analysis.

### Adoptive fat transplantation

Adoptive fat transplantation was performed in lipodystrophic mice and control littermates at a steady state or after one cycle of 1.5% DSS treatment. First, 500 to 600 mg of parametrial donor fat was harvested from sex-matched WT littermates or *ob/ob* mice. Second, lipodystrophic or control mice were anesthetized by isoflurane inhalation and an additional subcutaneous injection of tramadol (25 mg/kg, Grünenthal). Laparotomies were performed to introduce donor adipose tissue into the peritoneal cavity of recipient animals, as previously described^55^. After surgery, mice were treated with 0.2 mg/mL tramadol (Grünenthal) for three days. One month later, at sacrifice, the grafts were weighed and evaluated for attachment and vascularization using binocular surgical microscopy.

### Histopathologic analysis

Histopathologic analyses of colon sections from mice subjected to chronic DSS-induced colitis ± adoptive fat grafting were performed in a blinded fashion using an additive scoring system ranging from 0 to a maximum of 21 points for inflammation, as previously described^56^.

### Flow cytometry

For single cell analysis, spleens and mesenteric lymph nodes from lipodystrophic mice and control littermates were disassociated and filtered through a 70 μM cell strainer (Thermofisher Scientific) using MACS buffer consisting of PBS (Gibco) supplemented with 0.5% Fraction V bovine serum albumin (BSA) (Sigma-Aldrich). Cells were then resuspended in RBC lysis buffer (8.9 g NH_4_Cl, 1 g KHCO_3_, 0.038 g EDTA, 1 L distilled water) for 2 min at RT, then washed and filtered again through a 40 μM cell strainer (Thermofisher Scientific). For the terminal ileum and colon, tissue sections were opened longitudinally and cut into small pieces for two washes in 1 mM EDTA solution (Sigma-Aldrich) for 15 min at 37 °C under shaking conditions at 250 rpm. Tissue pieces were then digested in complete media supplemented with 200 U/mL Collagenase D (Roche) for 90 min under shaking conditions (250-300 rpm, 37 °C). Cells were then subjected to a 40/100% discontinuous Percoll gradient (GE Healthcare), 1200 g for 25 min at RT, to obtain mononuclear cell fractions. Finally, cell pellets were washed and resuspended for activation at 37 °C for 4 hours in RPMI medium 1640 (1X) (Gibco), supplemented with 10% fetal bovine serum (FBS) (Sigma-Aldrich), 10% penicillin-streptomycin (Thermofisher Scientific), 50 μM 2-mercaptoethanol (Gibco) with or without 20 ng/mL phorbol 12-myristate 13-acetate (PMA) (Sigma-Aldrich) and 1 μg/mL ionomycin (Sigma-Aldrich), and 10 μg/mL brefeldin A (Sigma-Aldrich) 2 hours before harvest. The cells were then washed and stained with viability dyes for 10 min at 4 °C. For surface staining, pellets were resuspended in MACS buffer and incubated with fluorescently labeled antibodies for 15 min on ice in the dark. Finally, cells were fixed in fixation/permeabilization buffer (eBioscience) according to the manufacturer’s protocol for 45 min at 4 °C. Pellets were washed twice in permeabilization buffer (eBioscience) and stained with antibody cocktails against intracellular molecules for 25 min at RT. A complete list of antibodies and viability dyes is provided in **Suppl. Table 5**. Samples were finally collected on a BD FACSCanto II device (BD). Data were analyzed using the FlowJo software version 10.6.2 (FlowJo).

### RNA-bulk seq

For RNA extraction, 1 cm of distal colon tissue, previously stored at −80 °C in RNA later, was first homogenized for 30 seconds at 50 Hz in lysis buffer (RLT buffer + β-mercaptoethanol) (Qiagen) supplemented with Reagent DX (Qiagen) using a TissueLyser LT Adapter. Second, homogenized tissue suspensions were processed following the Rneasy Mini protocol (Qiagen) according to the manufacturer’s instructions. Total RNA was sequenced on an Illumina platform. HISAT2^57^ was used to perform alignments to the Mus musculus reference genome assembly GRCm38 (mm10) after data quality control and filtering. After gene expression quantification, the differential gene expression analysis was performed using DESeq2 software, and significant differences were considered if |log2(FoldChange)| ≥ 1 & padj ≤ 0.05. Downstream pathway and gene enrichment analyses were performed using the g:Profiler platform or the GSEA 4.3.2 software^58^.

### ELISA

Serum was obtained from lipodystrophic mice and littermate WT controls. Serum levels of leptin or anti-E. coli (LPS) IgG were determined using the mouse Leptin DuoSet ELISA kit (R&D Systems) or mouse anti-*Escherichia* (*E.*) *coli* lipopolysaccharide (LPS) (O111:B4) Antibody ELISA Kit (Chondrex), respectively.

### Western blot analyses

All buffers used for immunoblotting are listed in **Suppl. Table 6**. For the analysis of tight junction proteins, distal colon sections obtained from lipodystrophic mice and WT littermates were mechanically disassociated in an ice-cold membrane lysis buffer using a FastPrep-24 homogenizer (MP Biomedical, Irvine, CA). After the first centrifugation (1000 g, 5 min, 4 °C), supernatants were once more centrifuged (42,000 g, 30 min, 4 °C) for membrane protein collection. The protein fraction was resolved in total lysis buffer and incubated 5 min at 95 °C in Laemmli buffer. Samples were loaded on a 5% stacking gel and then separated on a 12% SDS polyacrylamide gel and transferred onto a polyvinylidene fluoride (PVDF) membrane (#NBA085S001EA, Perkin Elmer, Rodgau, Germany). PVDF Membranes were blocked in blocking solution and incubated overnight with primary antibodies (**Suppl. Table 7**) at 4 °C on a shaker. Membranes were washed in 1X Tris-buffered saline with 0.1% Tween® 20 solution and incubated with the respective peroxidase-conjugated secondary antibodies (Dako, Denmark). Finally, membranes were washed, incubated in SuperSignalWest Pico PLUS buffer (#34580, ThermoFisher) and detected using a Fusion FX7 chemiluminescence imager (Vilber Lourmat). Densitometric analysis was performed using AIDA software (Elysia, North-Rhine Westphalia, Germany), and all values were normalized to the β-actin signal of the respective blot and then the mean of the WT controls.

### Calcium influx measurements in mice splenocytes

Spleens collected from lipodystrophic and WT mice were processed as mentioned above to obtain single cell suspension of splenocytes. Cells were then stained and collected using a FACSCanto II flow cytometer (BD) as previously described^16^. Briefly, splenocytes were stained with 2 μg/mL calcium-sensing dye Fluo-4 AM (ThermoFisher Scientific) for 30 min on ice followed by an anti-mouse antibody cocktail targeting surface markers for T cells (CD4, CD8) for 15 min on ice in the dark. Cells were then washed using PBS supplemented with 10% FBS before being resuspended in 0 mM Ca^2+^ Ringer’s solution (**Suppl. Table 8**). Baseline intracellular Ca^2+^ (*f_0_*) was then measured for 30 or 40 seconds. Cells were then stimulated with 1 μM thapsigargin (Merck), and after 300 seconds a final concentration of 2 mM Ca^2+^-containing Ringer’s solution was added. The sample was then acquired for another 120 seconds. The mean fluorescence intensity (MFI) of Fluo-4 (*f*) was normalized to the mean MFI detected during the 40-second baseline measurement (*f_0_*), and the resulting *f/f_0_* ratio was plotted against a time (t) axis. Graphs were generated using GraphPad Prism v.9 software (GraphPad).

## Notes

### Competing Interest Statement

B.S. received grant support by Pfizer, served as consultant for Abbvie, BMS, Boehringer, Endpoint Health, Falk, Galapagos, Gilead, Janssen, Johnson & Johnson, Landos, Lilly, MSD, Pfizer, Takeda (BS served as representative of the Charité) and received speaker's fees from Abbvie, AlfaSigma, BMS, CED Service GmbH, Falk, Ferring, Galapagos, Janssen, Lilly, Pfizer, Takeda (BS served as representative of the Charité). C.W. received grant support by Pfizer, served as consultant for Pfizer and received speaker's fees from Falk, Ferring, Janssen. The other authors declare to not have any competing interests.

### Author Declarations

Study was approved by the institutional review board of Charite - Universitatsmedizin Berlin (Approval number EA1/200/17). Written informed consent was obtained from all healthy volunteers and patients as approved by the institutional review board of Charite - Universitatsmedizin Berlin.

