## Supplemental tables and figures for "T cell-intrinsic lymphoproliferation as a key driver of intestinal autoimmunity in acquired generalized lipodystrophy"

### Supplementary Materials Letizia et al.

#### Supplementary tables

##### Suppl. Table 1. Cohort characteristics

| *Patients´ characteristics* Total Patients 11 | Crohn´s disease | AGLCD | Non-IBD |
|---|---|---|---|
| Patients number (%) | 6 (54.55%) | 1 (9.09%) | 4 (36.36%) |
| Mean age in years (range) | 55 (39-71) | (25-35) | 73.33 (69-77) |
| Males number (%) | 1 (16.67%) | 1 (100.00%) | 3 (75.00%) |
| % of current or prior aminosalicylates medication (AZA) | 16.67% | 100.00% | 0.00% |
| Current or prior corticosteroids medication (%) | 50.00% | 100.00% | 0.00% |
| Current or prior immunomodulators medication (%) | 0.00% | 100.00% | 0.00% |
| Current or prior biologic therapies (agent used) (%) | 16.67% (ADA) | 100.00% (IFX) | 0.00% |
| Chemotherapy at inclusion (%) | 0.00% | 0.00% | 25.00% |
| Mean leukocyte count per nl at inclusion (range) | 5.69 (4.01-9.52) | --- | 7.11 (5.77-8.47) |

#### Suppl. Table 2. Cell culture and in vitro assays medium

| Complete Medium | Cat. No. | Company |
|---|---|---|
| RPMI Medium 1640 (1X) | #11875093 | Gibco |
| 10% Fetal bovine serum (FBS) | --- | Sigma-Aldrich |
| 10% Penicillin-Streptomycin | #15140122 | Thermofisher Scientific |
| **Activation Medium** | --- | --- |
| RPMI Medium 1640 (1X) | #11875093 | Gibco |
| 10% FBS | --- | Sigma-Aldrich |
| 10% Penicillin-Streptomycin | #15140122 | Thermofisher Scientific |
| 50 µM 2-Mercaptoethanol | #31350010 | Gibco |
| 20 ng/ml phorbol 12-myristate 13-acetate (PMA) | #P8139 | Sigma-Aldrich |
| 1 µg/ml ionomycin | #I0634 | Sigma-Aldrich |
| **T cell expansion Medium** | --- | --- |
| RPMI Medium 1640 (1X) | #11875093 | Gibco |
| 10% FBS | --- | Sigma-Aldrich |
| 10% Penicillin-Streptomycin | #15140122 | Thermofisher Scientific |
| 50 µM 2-Mercaptoethanol | #31350010 | Gibco |
| 250 U/ml human recombinant IL-2 | #200-02 | PeproTech, Rocky Hills, New |
| 1:100 or 1:500 T Cell TransAct™, human | #130-111-160 | Miltenyi Biotec, NRW, Germany |
| **Human LPMC digestion Medium** | --- | --- |
| RPMI Medium 1640 (1X) | #11875093 | Gibco |
| 10% FBS | --- | Sigma-Aldrich |
| 10% Penicillin-Streptomycin | #15140122 | Thermofisher Scientific |
| 0.15 mg/ml collagenase A | #11088793001 | Roche, Basel, Switzerland |
| **Murine LPMC digestion Medium** | --- | --- |
| RPMI Medium 1640 (1X) | #11875093 | Gibco |
| 10% FBS | --- | Sigma-Aldrich |
| 10% Penicillin-Streptomycin | #15140122 | Thermofisher Scientific |
| 200 U/ml collagenase D | #11088858001 | Roche |

#### Suppl. Table 3: Anti-human antibodies used in mass cytometry

| Metal | Target | Clone | Cat. No./ Company | Dilution |
|---|---|---|---|---|
| 89Y | CD45 | HI30 | #3089003B / Fluidigm | 1:100 |
| 141Pr | CD45 | HI30 | #3141009B / Fluidigm | 1:100 |
| 142Nd | CD19 | HIB19 | #3142001B / Fluidigm | 1:100 |
| 143Nd | CD45RA | HI100 | #3143006B / Fluidigm | 1:100 |
| 144Nd | IL-4 | MP4-25D2 | #3144010B / Fluidigm | 1:100 |
| 145Nd | CD4 | RPA-T4 | #3145001B / Fluidigm | 1:50 |
| 146Nd | TNFα | Mab11 | #3146010B / Fluidigm | 1:100 |
| 147Sm | CD11c | Bu15 | #3147008B / Fluidigm | 1:200 |
| 148Nd | IgA | Polyclonal | #3148007B / Fluidigm | 1:100 |
| 149Sm | CD25 | 2A3 | #3149010B / Fluidigm | 1:100 |
| 150 Nd | CD86 | IT2.2 | #3150020B / Fluidigm | 1:100 |
| 151Eu | CD103 | Ber-ACT8 | #3151011B / Fluidigm | 1:100 |
| 152Sm | CD95/Fas | DX2 | #3152017B / Fluidigm | 1:200 |
| 153Eu | IgM | MHM-88 | #314502 / Biolegend | 1:200 |
| 154Sm | CD3 | UCTH1 | #3154003B / Fluidigm | 1:100 |
| 155Gd | CD56 | B159 | #3155008B / Fluidigm | 1:100 |
| 156Gd | IL-6 | MQ2-13AS | #3156011B / Fluidigm | 1:100 |
| 158Gd | IFNγ | B27 | #3158017 / Fluidigm | 1:400 |
| 159Tb | CCR7 | G043H7 | #3159003C / Fluidigm | 1:200 |
| 160Gd | CD27 | O323 | #302802 / Biolegend | 1:200 |
| 161Dy | IL-10 | JES3-9D7 | #3161008B / Fluidigm | 1:100 |
| 161Dy | IL-23p19 | 23dcdp | #3161010B / Fluidigm | 1:100 |
| 162Dy | CD8 | RPA-T8 | #3162015B / Fluidigm | 1:100 |
| 163Dy | CD33 | WM53 | #3163023B / Fluidigm | 1:100 |
| 163Dy | Granzyme B | QA16A02 | #3173006B / Fluidigm | 1:100 |
| 164Dy | CD45RO | UCHL1 | #3164007B / Fluidigm | 1:100 |
| 165Ho | CD40 | 5C3 | #3165005B / Fluidigm | 1:100 |
| 166Er | IL-2 | MQ117H12 | #3166002B / Fluidigm | 1:100 |
| 167Er | CD38 | HIT2 | #3167001B / Fluidigm | 1:200 |
| 168Er | CD40L | 24-31 | #3168006B / Fluidigm | 1:100 |
| 169Tm | IL-13 | JES105A2 | #3169016 / Fluidigm | 1:100 |
| 170Er | CD137 | 4B4-1 | #3209015B / Biolegend | 1:100 |
| 171Yb | CD178/FasL | NOK-1 | #306402 / Biolegend | 1:100 |
| 172Yb | IL-17 | BL168 | #3172020B / Fluidigm | 1:100 |
| 173Yb | HLA-DR | L243 | #3173005B / Fluidigm | 1:200 |
| 174Yb | PD-1 | EH12.2H7 | #3174020B / Fluidigm | 1:200 |
| 175Lu | CD14 | M5E2 | #3175015B / Fluidigm | 1:50 |
| 175Lu | Perforin | B-D48 | #3175004B / Fluidigm | 1:100 |
| 176Yb | IL-7R | A019D5 | #3176004B / Fluidigm | 1:50 |
| 209Bi | CD11b | ICRF44 | #3209003B / Fluidigm | 1:100 |

#### Suppl. Table 4. Anti-human antibodies flow cytometry

| Fluorochrome | Target | Clone | Cat. No. / Company | Dilution |
|---|---|---|---|---|
| FITC | CD14 | 61D3 | #11-0149-42 / eBioscience | 1:50 |
| FITC | CD19 | HIB19 | #11-0199-42 / eBioscience | 1:100 |
| Pe-Cy7 | CD16 | 3G8 | #560918 / BD | 1:200 |
| APC | CD14 | 63D3 | #367118 / Biolegend | 1:50 |
| APC | CD4 | RPA-T4 | #555349 / BD | 1:20 |
| APC | CD3 | OKT3 | #17-0037-42 / eBioscience | 1:100 |
| APC | CD56 | TULY56 | #17-0566-42 / eBioscience | 1:200 |
| APC-Cy7 | CD19 | HIB-19 | #302218 / Biolegend | 1:100 |
| APC-Cy7 | CD8 | SK1 | #344713 / Biolegend | 1:100 |
| Viogreen | CD3 | REA613 | #130-113-142 / Miltenyi Biotec | 1:00 |
| BV510 | CD4 | RPA-TA | #300546 / Biolegend | 1:20 |

#### Table 5. Anti-mouse antibodies for flow cytometry

| Fluorochrome | Target | Clone | Cat. No. / Company | Dilution |
|---|---|---|---|---|
| FITC | CD4 | GK1.5 | #100406 / Biolegend | 1:100 |
| A488 | Eomes | Dan11mag | #53-4875-80 / eBioscience | 1:100 |
| PE | Foxp3 | FJK-16s | #12-5773-82 / eBioscience | 1:100 |
| PE | NK1.1 | PK136 | #12-5941-81 / eBioscience | 1:200 |
| PE | CD49a | REA493 | #130-124-706 / Miltenyi Biotec | 1:50 |
| PerCP-Cy5.5 | IL-17A | TC11-18H10.1 | #506920 / Biolegend | 1:200 |
| PerCP-Cy5.5 | CD3 | 145-2C11 | #551163 / BD | 1:200 |
| PerCP-Cy5.5 | CD27 | LG.3A10 | #124214 / BD | 1:100 |
| PerCP-Cy5.5 | NK1.1 | PK136 | #108728 / Biolegend | 1:50 |
| Pe-Cy7 | TNFα | MP6-XT22 | #557644 / BD | 1:200 |
| Pe-Cy7 | F4/80 | BM8 | #123118 / Biolegend | 1:300 |
| Pe-Cy7 | CD49b | DX-5 | #12-5971-82 / eBioscience | 1:50 |
| APC | IFNγ | XMG1.2 | #554413 / BD | 1:200 |
| APC | T-bet | 4B10 | #644814 / Biolegend | 1:500 |
| APC-Cy7 | CD4 | GK1.5 | # 100414 / Biolegend | 1:500 |
| APC-Cy7 | CD45 | 30-F11 | #103116 / Biolegend | 1:200 |
| APC-Cy7 | CD11b | M1/70 | #101226 / Biolegend | 1:100 |
| APC-Cy7 | GR-1 | RB6-8C5 | #108434 / BD | 1:200 |
| APC-Cy7 | CD19 | 6D5 | #115530 / Biolegend | 1:50 |
| APC-Cy7 | F4/80 | BM8 | #123118 / Biolegend | 1:200 |
| APC-Cy7 | CD3e | 145-2C11 | #557596 / BD | 1:50 |
| APC-Cy7 | FcERIa | MAR-1 | #134326 / Biolegend | 1:100 |
| eFluor450 | CD8a | 53-6.7 | #48-0081-82 / eBioscience | 1:200 |
| BV421 | GR-1 | RB6-8C5 | #108434 / Biolegend | 1:200 |
| BV421 | CD11b | M1/70 | #101236 / Biolegend | 1:100 |
| V500 | CD45 | 30-F11 | #561487 / BD | 1:200 |
| Zombie Violet | Viability Dye | | #423114 / Biolegend | 1:1000 |
| Zombie Aqua | Viability Dye | | #423102 / Biolegend | 1:1000 |
| eFluor780 | Viability Dye | | #65-0865-14 / eBioscience | 1:1000 |

#### Suppl.Table 6. Buffers used in western blotting

| **Tissue lysis buffer** | **Catalog number (Cat. No.)** | **Company** |
|---|---|---|
| 1 M Tris-HCl pH 7.4 | #9090.3 | Carl Roth, Karlsruhe, Germany |
| 1 M $MgCl_2$ | #5833 | Merck, Darmstadt, Germany |
| 0.5 M ethylenediaminetetraacetic acid (EDTA) | #E6511 | Sigma-Aldrich, St. Louis, Missouri |
| 0.5 M ethylene glycol-bis (β-aminoethyl ether)-N,N,N′,N′-tetraacetic acid (EGTA) | #324626 | Sigma-Aldrich |
| Protease inhibitor cocktail | #P8340 | Sigma-Aldrich |
| **RIPA buffer cocktail** | --- | --- |
| 1X RIPA Buffer | #R0278 | Merck |
| 14X Protease inhibitor cocktail I | #20-201 | Merck |
| 10X Phosphatase inhibitor cocktail | #P5726 | Merck |
| 200mM $NaVO_4$ | #S6508 | Merck |
| 200mM NaF | #106449 | Merck |
| **8% Sodium dodecyl sulfate (SDS) running gel** | --- | --- |
| 40% acrylamide mix | #1610140 | Biorad |
| 5 mM Tris base pH 8.8 | #4855.2 | Carl Roth |
| 10% SDS | #2326.2 | Carl Roth |
| 10% ammonium persulfate (APS) | #7727-54-0 | Sigma-Aldrich |
| 1% tetramethylethylenediamine (TEMED) | #161-0800 | Biorad |
| **5% Stacking gel** | --- | --- |
| 40% acrylamide mix | #1610140 | Biorad |
| 1.0 M Tris base pH 6.8 | #4855.2 | Carl Roth |
| 10% SDS | #2326.2 | Carl Roth |
| 10% APS | #7727-54-0 | Sigma-Aldrich |
| 1% TEMED | #161-0800 | Biorad |
| **Laemmli buffer (6X)** | --- | --- |
| 60 mM Tris-HCl pH 6.8 | #9090.3 | Carl Roth |
| 12% SDS | #2326.2 | Carl Roth |
| 47% glycerol | #3783.1 | Carl Roth |
| 0.06% bromophenol blue | #8122 | Merck |
| 12.5% β-mercaptoethanol | #60-24-2 | Sigma-Aldrich |
| **Electrophoresis buffer (10X)** | --- | --- |
| 144 g glycine | #3908.3 | Carl Roth |
| 30 g Tris base | #4855.5 | Carl Roth |
| 10 g SDS | #2326.2 | Carl Roth |
| 1 L distilled water | --- | --- |
| **Transfer buffer (10X)** | --- | --- |
| 30.3 g Tris base | #4855.5 | Carl Roth |
| 144 g glycine | #3908.3 | Carl Roth |
| 1 L distilled water | --- | --- |
| **Transfer buffer (1X)** | --- | --- |
| 100 ml transfer buffer (10X) | --- | --- |
| 200 ml 100% methanol | #32213 | Merck |
| 700 ml distilled water | --- | --- |
| **Tris-Buffered Saline (TBS) pH 7.6 (10X)** | --- | --- |
| 24 g Tris base | #4855.2 | Carl Roth |
| 88 g NaCl | #9265.2 | Carl Roth |

| | | |
|---|---|---|
| 1 L distilled water | --- | --- |
| **Tris-Buffered Saline-Tween20 (TBS-T) (1X)** | --- | --- |
| 100 ml TBS pH 7.6 (10X) | --- | --- |
| 1 ml Tween | #9127.1 | Carl Roth |
| 900 ml distilled water | --- | --- |
| **5% Blocking buffer** | --- | --- |
| 2.5 g bovine serum albumin (BSA) (Fraction V) | #9048-46-8 | Sigma-Aldrich |
| 50 ml TBS-T (1X) | --- | --- |
| 2.5 mM L-glutamine | #56-85-9 | Sigma-Aldrich |

#### Suppl. Table 7. Antibodies used for western blot

| Target | Clone | Cat. No. / Company | Dilution |
|---|---|---|---|
| CLDN1 | Polyclonal | #19000 / Invitrogen | 1:1000 |
| CLDN2 | MH44 | #16100 / Invitrogen | 1:1000 |
| CLDN3 | Polyclonal | #41700 / Invitrogen | 1:1000 |
| CLDN4 | 3E2C1 | #29400 / Invitrogen | 1:1000 |
| CLDN7 | Polyclonal | #49100 / Invitrogen | 1:1000 |
| OCCL | Polyclonal | #711500 / Invitrogen | 1:1000 |
| TRIC | 54H19L38 | #700191 / Invitrogen | 1:1000 |
| MD3 | Polyclonal | #5567-1-AP/ Proteintech, Rosemont, IL | 1:1000 |
| ZO-1 | ZO1-1A12 | #33-9100 / Invitrogen | 1:1000 |
| LSR | Polyclonal | #HPA007270 / Atlas antibodies | 1:1000 |
| ILDR1 | Polyclonal | #bs-11013R / Thermofisher | 1:1000 |
| ACTB | AC-15 | #A5441 / Sigma-Aldrich | 1:10.000 |

#### Suppl. Table 8. Buffers used for $Ca^{2+}$ influx

| Ringer Solution | Cat. No. | Company |
|---|---|---|
| 155 mM NaCl | #9265.2 | Merck |
| 4.5 mM KCl | #7447-40-7 | Merck |
| 3 mM $MgCl_2$ | #5833 | Merck |
| 10 mM D-glucose | #X997.2 | Merck |
| 5 mM Na-HEPES | #75277-39-3 | Merck |
| ± 4 mM $CaCl_2$ | #2.382 | Merck |

#### Suppl. Table 9. Top 20 expanded T cell clonotypes in the AGLCD patient

| ID | CTaa | n |
|---|---|---|
| 1 | CVVSVFSGGYNKLIF_CASAAGGNSHNYPSGANVLTF | 156 |
| 2 | CVVNISYSGYALNF_CASAAGGNSHNYPSGANVLTF | 131 |
| 3 | CVVNISYSGYALNF;CVVSVFSGGYNKLIF_CASAAGGNSHNYPSGANVLTF | 83 |
| 4 | NA_CASAAGGNSHNYPSGANVLTF | 76 |
| 5 | NA_CSARGGLNSPLHF | 72 |
| 6 | NA_CASSLEGYGYTF | 63 |
| 7 | CGTDTGNQFYF_CASSLWGPRSNQPQHF | 59 |

| | | |
|---|---|---|
| 8 | CAGLSNTGNQFYF_CASSLEGYGYTF | 54 |
| 9 | NA_CASSFYPGRSGANVLTF | 52 |
| 10 | CAGQPPASGAGSYQLTF_CSARGGLNSPLHF | 43 |
| 11 | CAETFSGGYNKLIF_CASSSWGQGYGYTF | 41 |
| 12 | CAMRLTDSWGKLQF_CASSLEVGYEAFF | 36 |
| 13 | NA_CSARDRPGRVLYTDTQYF | 32 |
| 14 | CAGLADSGGGADGLTF_CASSFYPGRSGANVLTF | 30 |
| 15 | CIVRGGTGNQFYF_CASSQRNLDGYNSPLHF | 26 |
| 16 | CALVSGGYQKVTF_CASSLTGADTGELFF | 24 |
| 17 | CVVTRPSGGYNKLIF_CASSQELAGGKIPLSSYNEQFF | 24 |
| 18 | CAETFSGGYNKLIF_NA | 23 |
| 19 | CAVEGIKAAGNKLTF_CASSEGSYGYTF | 22 |
| 20 | CAVILFSGGYNKLIF_CSARDRPGRVLYTDTQYF | 19 |

### Supplementary Figure 1

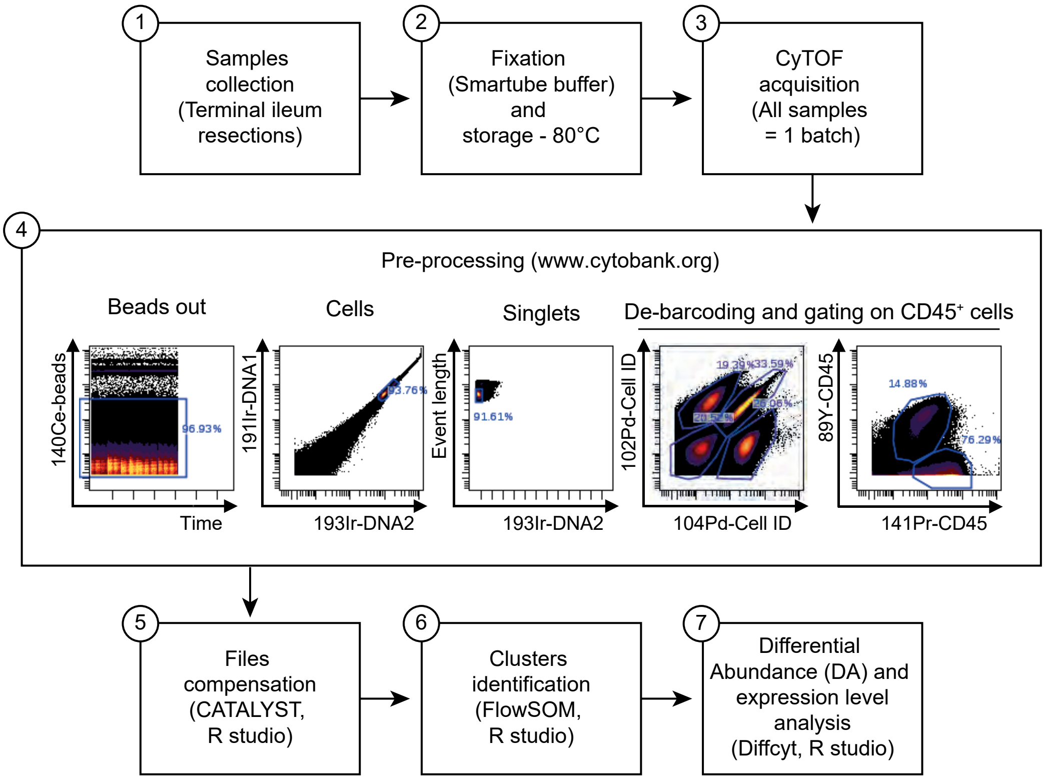


**Suppl. Figure 1. Experimental setup to characterize gut-resident immune cells via mass cytometry.** Lamina propria mononuclear cells (LPMCs) were isolated from terminal ileum samples obtained from 6 Crohn's disease (CD) patients, 4 non-IBD control patients and one AGLCD patient. Cells were stored at -80°C until acquisition. All the samples collected were acquired on a CyTOF2 mass cytometer in one batch. For data analysis, resulting flow cytometry standard (FCS) files were first normalized and then uploaded to Cytobank (www.cytobank.org) for gating of single, live cells and de-barcoding. Individual FCS files were compensated using the R package CATALYST and 17 clusters were identified using the R package FlowSOM. Differential abundance and expression level analyses were then done using the R package Diffyct.

Supplementary Figure 2

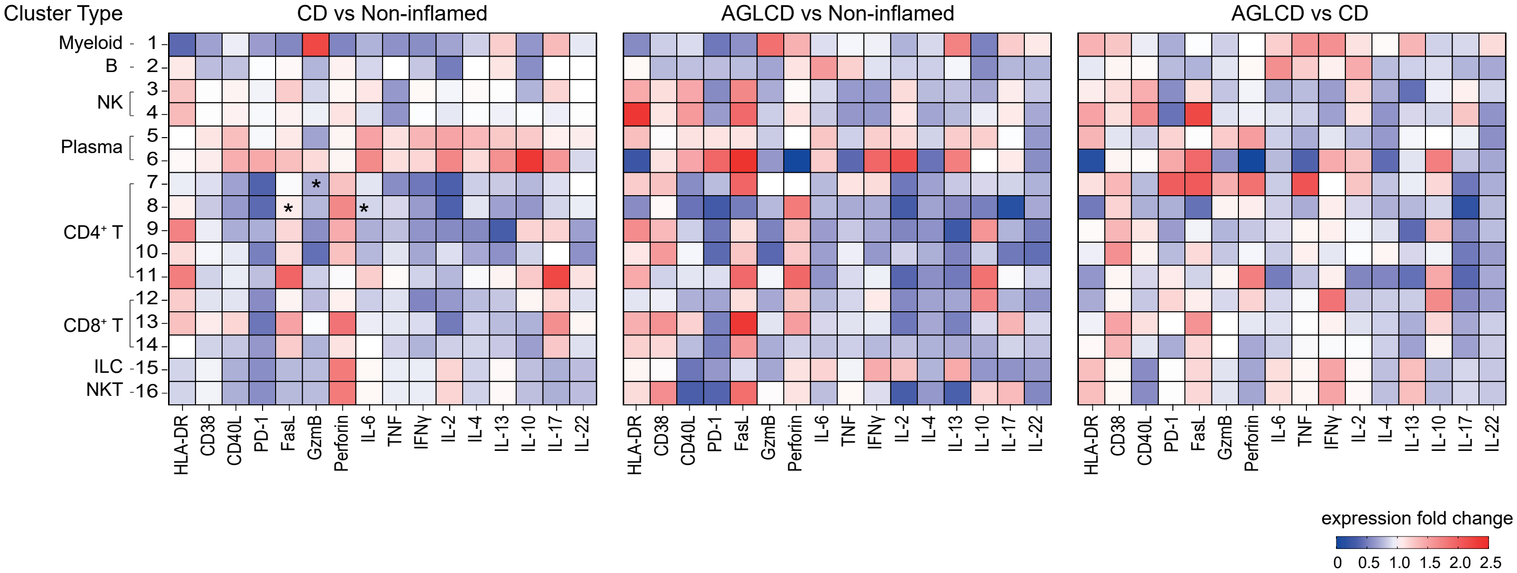


**Suppl. Figure 2. Differences in expression levels of activation markers, cytokines and pore-forming cytolytic proteins between AGLCD, CD, and non-inflamed control cell clusters.** LPMCs were isolated from AGLCD patient and controls (AGLCD: n=1, CD: n= 6, non-inflamed: n=4) and then activated for 4 h in vitro with PMA/ionomycin. Heatmaps show the median fold change of cytokines and activation marker expression in different immune cell populations. Significant differences were calculated using a paired Wilcoxon matched-pairs signed rank test, *P < 0.05.

Supplementary Figure 3

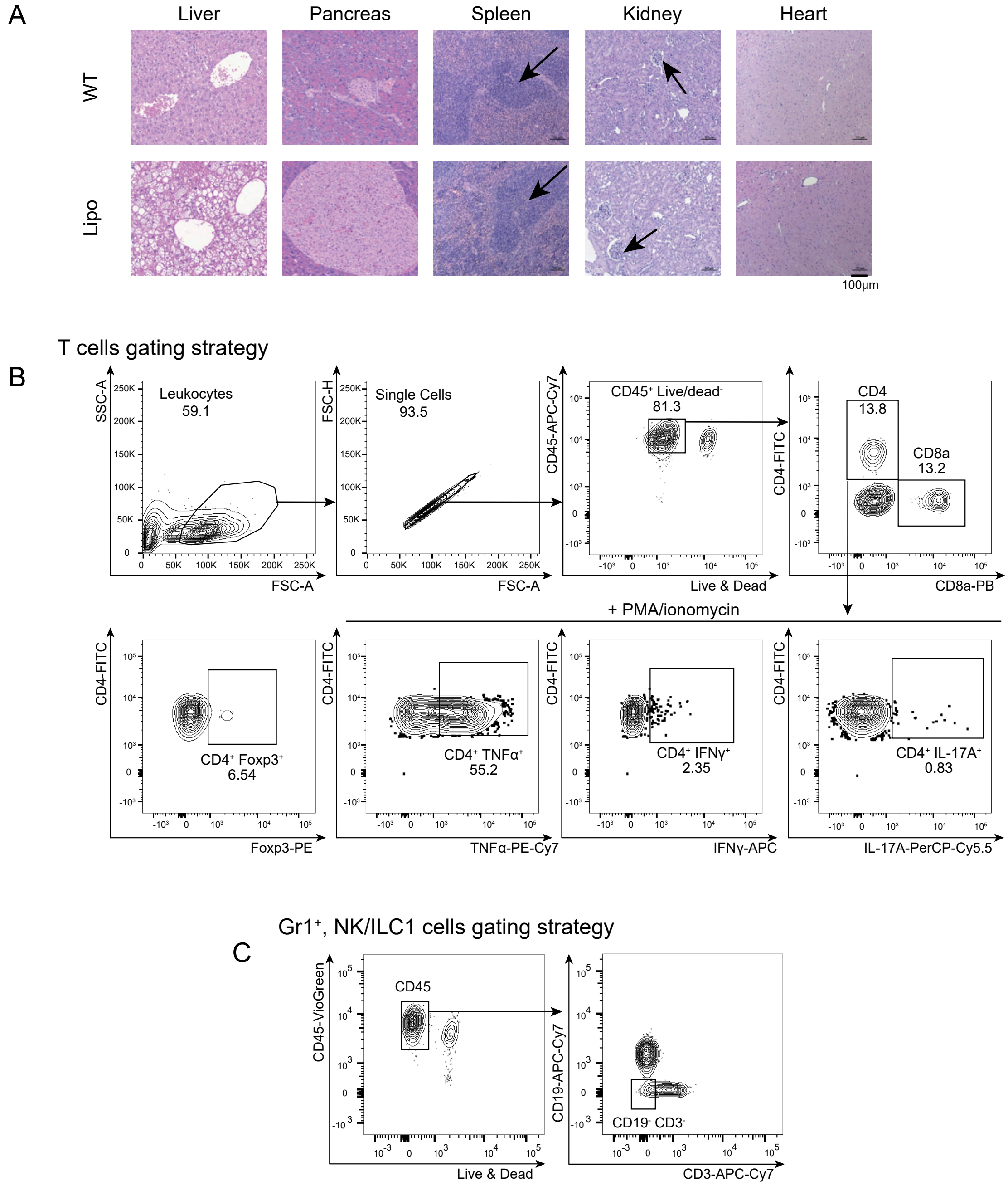


**Suppl. Figure 3. Representative flow cytometry gating strategy for T cells and NK/ILC1 cells.** (A) Representative histologic analyses of various organs obtained from wild-type (WT) and lipodystrophic mice. Arrows point to the enlarged red pulp in the spleen and enlarged glomeruli in the kidney. (B) Immune cells isolated from spleen or intestinal tissues of WT and lipodystrophic mice were characterized using flow cytometry. Cells were gated for scattering (SSC-A/FCS-A), then doublets (FSC-H/FSC-A) were excluded, and only live immune (CD45+ Live&Dead-) cells were selected for further evaluation. For cytokine staining of CD4+ or CD8+ T cells, cells were stimulated in vitro for 4 h with PMA/ionomycin. (C) For NK/ILC1 cell populations, only live immune (CD45+ Live&Dead-) cells that were also CD19-CD3- were selected for further evaluation.

Supplementary Figure 4

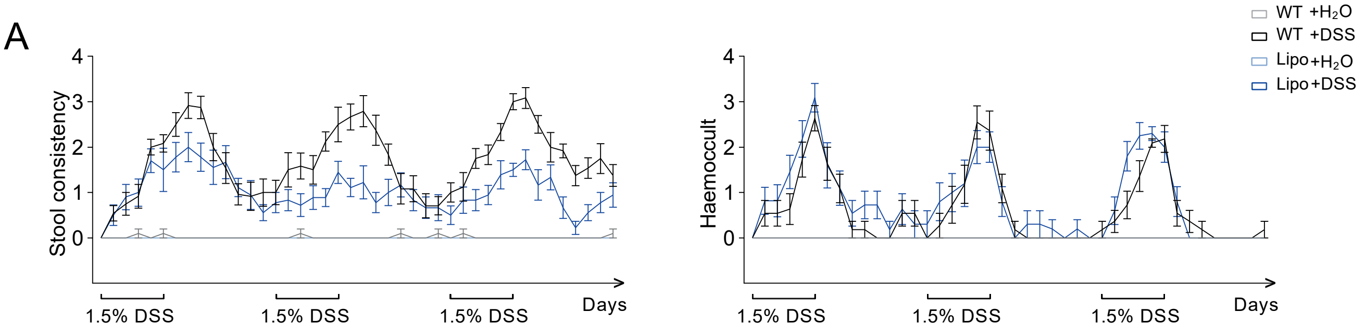


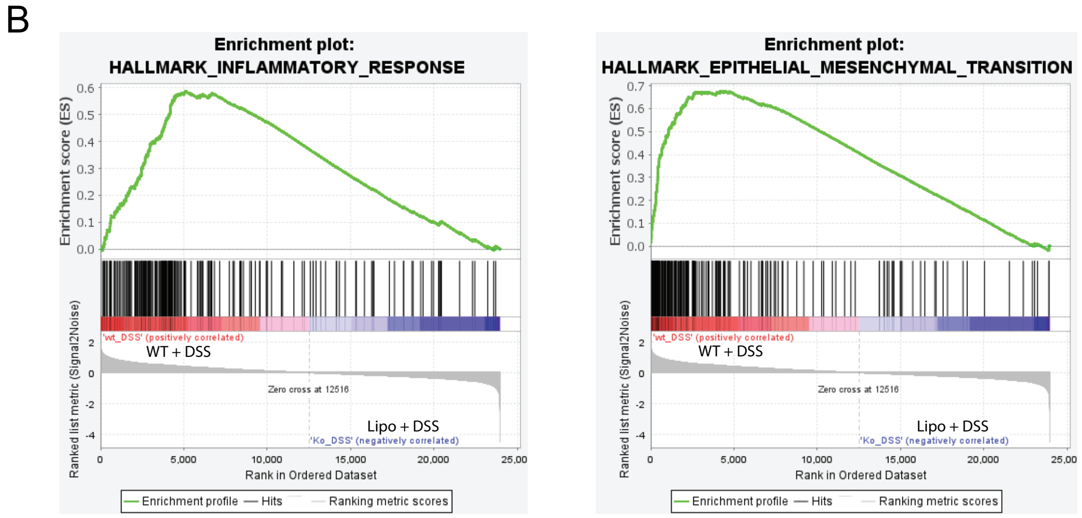


**Suppl. Figure 4. Lipodystrophic mice have less severe colitis and altered immune responses compared to WT mice in the chronic DSS model.** Mice received three cycles of 1.5% DSS and were monitored for (A) stool consistency and tested for hemoccult blood during the experiment. Data was pooled from two independent experiments. Lines are projections of mean values, and the error bars represent the standard error mean. (WT+ H2O: n=10; WT+ DSS: n=11; Lipo + H2O: n=6; Lipo + DSS: n=11). (B) Gene set enrichment analysis (GSEA) plot showing upregulation of genes involved in the inflammatory response and epithelial-mesenchymal transition gene sets in the WT DSS-treated mice compared to lipoatrophic mice DSS-treated mice (n = 5 per group).

### Supplementary Figure 5

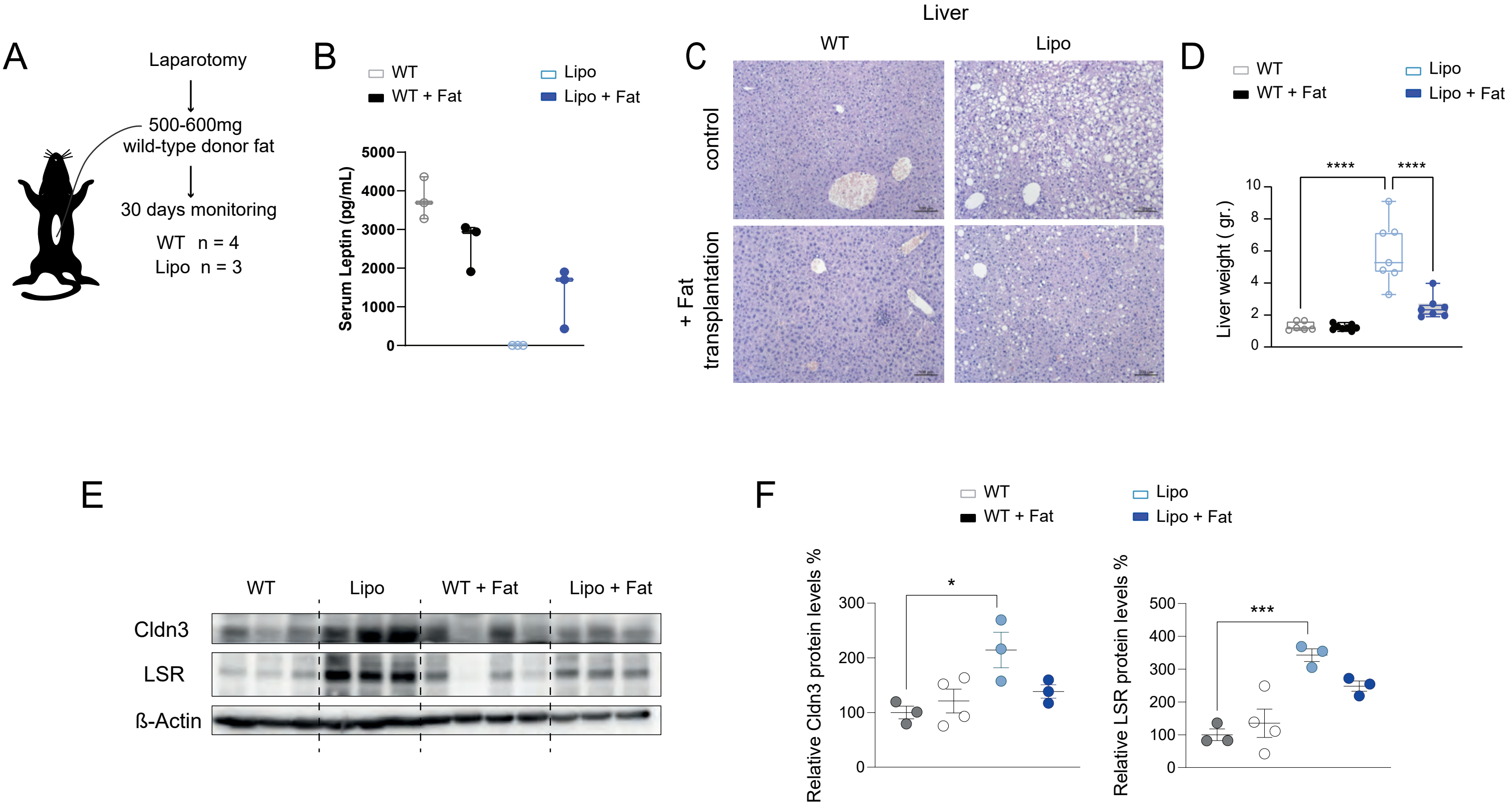


**Suppl. Figure 5. Allogenic fat transplantation reverses steatohepatitis and restores tight junction protein expression in lipodystrophic mice.** (A) Experimental design: lipodystrophic (Lipo) or wild-type (WT) mice were transplanted with 500-600 mg adipose tissue obtained from WT donor mice by performing mini-laparotomy. Mice were monitored for 30 days before organs were harvested for further experimentation. (B) Serum levels of leptin. (C) Representative H&E staining of liver sections, (D) liver weight, (E) Western blot showing expression levels of TJs proteins isolated from the colon lamina propria of mice (WT: n = 3; WT + fat: n = 4; Lipo: n = 3; Lipo + fat: n = 3). (F) Whisker and box plots show relative expression of Claudin 3 (Cldn3) and Lipoprotein receptor (LSR) proteins in lipoatrophic mice normalized to wild-type controls (wild-type = 100%). Boxes range from the 25th to the 75th percentile. Whisker plots show the minimum (smallest) and maximum (largest) values. The line in the box indicates the median. (SEM). Statistic was calculated by one-way ANOVA test.

#### Supplementary Figure 6

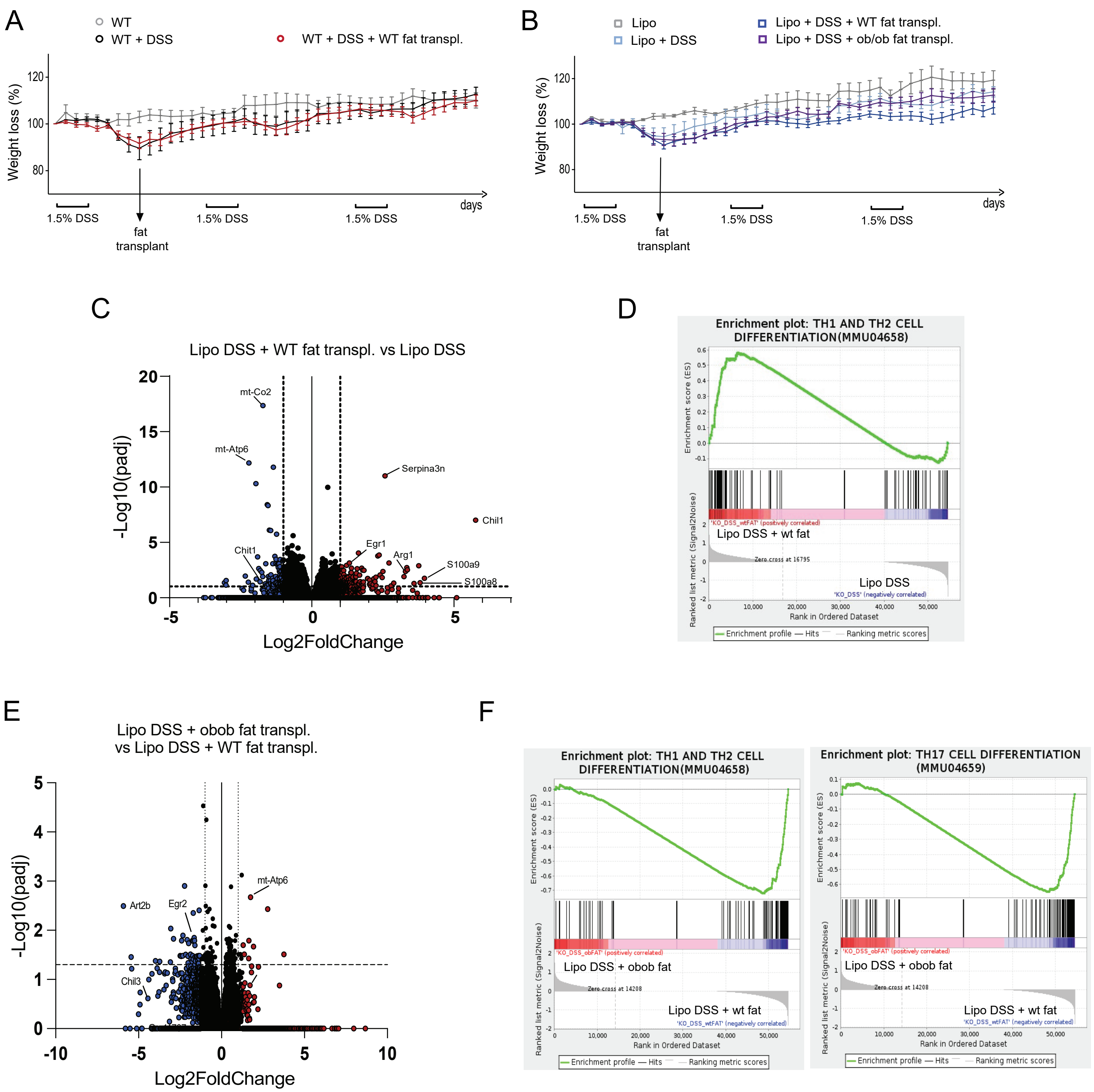


**Suppl. Figure 6. Lipodystrophic mice that received allogenic wild-type (WT) fat show reduced colitis compared to lipodystrophic mice that received allogenic ob/ob fat in the chronic DSS model**. Lipodystrophic (Lipo) or WT mice received 1 cycle of 1.5% DSS followed by transplantation with 500-600 mg adipose tissue obtained from WT or leptin-deficient ob/ob donor mice by performing mini-laparotomy. Seven days after transplantation, mice were subsequently challenged with 2 cycles of DSS treatment. Mice were monitored throughout the experiment and relative weight loss was recorded (A) for Lipo mice groups and (B) WT mice groups. Lines are projections of mean values. Data shown is pooled from 2 independent transplantation experiments (n=4-13). (C) Volcano plot showing differentially regulated genes between DSS-treated Lipo mice that received WT fat and DSS-treated Lipo mice (n=6 per group). (D) Gene set enrichment analysis (GSEA) plot showing upregulation of genes involved in Th1 and Th2 cell differentiation in DSS-treated lipodystrophic that received WT fat and DSS-treated Lipo mice. (E) Volcano plot showing differently regulated genes between DSS-treated lipodystrophic that received ob/ob fat and DSS-treated Lipo mice that received WT fat (n=4-6). (F) GSEA plot showing upregulation of genes involved in Th1, Th2 and Th17 cell differentiation in DSS-treated lipodystrophic that received WT fat and DSS-treated lipodystrophic that received ob/ob fat.
